# A mathematical model to identify optimal combinations of drug targets for dupilumab poor responders in atopic dermatitis

**DOI:** 10.1101/2021.02.08.21251317

**Authors:** Takuya Miyano, Alan D Irvine, Reiko J Tanaka

**Affiliations:** Department of Bioengineering, Imperial College London, UK; Pediatric Dermatology, Children’s Health Ireland at Crumlin, Dublin; Clinical Medicine, Trinity College Dublin, Dublin, Ireland

**Keywords:** atopic dermatitis, dupilumab, model-based meta-analysis, poor responders, quantitative systems pharmacology

## Abstract

**Background:** Several biologics for atopic dermatitis (AD) have demonstrated good efficacy in clinical trials, but with a substantial proportion of patients being identified as poor responders. This study aims to understand the pathophysiological backgrounds of patient variability in drug response, especially for dupilumab, and to identify promising drug targets in dupilumab poor responders.

**Methods:** We conducted model-based meta-analysis of recent clinical trials of AD biologics and developed a mathematical model that reproduces reported clinical efficacies for nine biological drugs (dupilumab, lebrikizumab, tralokinumab, secukinumab, fezakinumab, nemolizumab, tezepelumab, GBR 830, and recombinant interferon-gamma) by describing systems-level AD pathogenesis. Using this model, we simulated the clinical efficacy of hypothetical therapies on virtual patients.

**Results:** Our model reproduced reported time courses of %improved EASI and EASI-75 of the nine drugs. The global sensitivity analysis and model simulation indicated the baseline level of IL-13 could stratify dupilumab good responders. Model simulation on the efficacies of hypothetical therapies revealed that simultaneous inhibition of IL-13 and IL-22 was effective, whereas application of the nine biologic drugs was ineffective, for dupilumab poor responders (EASI-75 at 24 weeks: 21.6% vs. max. 1.9%).

**Conclusion:** Our model identified IL-13 as a potential predictive biomarker to stratify dupilumab good responders, and simultaneous inhibition of IL-13 and IL-22 as a promising drug therapy for dupilumab poor responders. This model will serve as a computational platform for model-informed drug development for precision medicine, as it allows evaluation of the effects of new potential drug targets and the mechanisms behind patient variability in drug response.

## 1. INTRODUCTION

Atopic dermatitis (AD) is the most common inflammatory skin disease, whose incidence is increasing in many areas of the world, especially urbanized areas, with a current worldwide prevalence of 5%-25%^1^. Primary symptoms of AD are relapsing pruritus and skin pain, impairing patients’ quality of life, for example by sleep disturbance and decreased work productivity especially in moderate-to-severe cases^2^. The pathogenesis of AD involves epidermal barriers abnormalities, dysbiosis and heterogeneous immunological dysregulations^3,4,5^ characterized by a dominant Type 2 immune activation including the T helper (Th) 2 axis and, depending on the lesional stage and ethnicity, varying degrees of upregulation of the Th1, Th17, and Th22 axes. These Th cells produce inflammatory cytokines such as interleukin (IL)-4, IL-13, IL-17A, IL-22, and IL-31, all of which have been identified and investigated as therapeutic targets for AD^4,5^.

Dupilumab, a monoclonal antibody that inhibits signaling from IL-4 and IL-13 by blocking their common IL-4 receptor subunit α (IL-4Rα), was approved as the first and, so far, the only AD-specific biologic in 2017^6^ for its promising efficacy demonstrated in clinical trials. The high efficacy of dupilumab confirmed the clinical validity of IL-4 and IL-13 as therapeutic targets for AD. However, dupilumab treatment was not effective for a sub-population of patients; the responder rates for dupilumab remain 44%-69% for Eczema Area and Severity Index (EASI)-75 (75% reduction in the EASI score^7, 8^) and 36%-39% for achievement of clear or almost clear skin in Investigator’s Global Assessment, respectively^9, 10^.

A significant percentage of poor responders was also observed for investigational drugs with other mechanisms of action (MoA), even if their clinical efficacy was confirmed for the study population average. It is thus of high clinical importance to identify underlying pathogenesis that causes the patients’ variability in responsiveness to each drug, and to investigate whether there are alternative drug targets for those poor responders.

The clinical efficacy of investigational drugs for AD have been evaluated in many clinical trials. Reviewing the combined results from the clinical trials, for example by network meta-analysis^11^, suggested hypothetical AD pathogenesis described as diagrammatic pathways^12,13^. However, such a qualitative framework does not account for patient stratification and for development of drugs that can be effective for poor responders to existing drugs, because underlying mechanisms for the variability in individual patients’ responsiveness cannot be evaluated. In addition, a simple correlation analysis between clinical efficacy and biological factors (e.g., transcriptome data) ^14, 15, 16^ is not suitable to identify appropriate biomarkers for patient stratification, as it may detect pseudo-correlations rather than the actual causal relationship. Instead, a quantitative model-based framework that describes AD pathogenesis is required to identify biomarkers for patient stratification and to evaluate the clinical efficacy of new hypothetical therapies such as the one that inhibits a combination of cytokines^17^.

As a quantitative approach to elucidate disease pathogenesis, quantitative systems pharmacology (QSP) has been successfully applied to enhance understanding of the pathogenesis of many inflammatory diseases in the context of translational drug development^18, 19^. QSP uses mathematical models to describe a systems-level understanding of pathogenesis and drug effects by integrating biological and pharmacological knowledge^20^. For example, the QSP approach has been applied to evaluate the efficacy of approved and experimental drugs in rheumatoid arthritis^21^, the effects of a hypothetical drug on cytokine behaviors in Crohn’s disease^22^, and the relationship between Th1/Th2 responses and exposure levels of lipopolysaccharide in asthma^23^. QSP modeling can be leveraged with model-based meta-analysis, which integrates data from different clinical trials with current understanding on disease pathogenesis, to make maximal use of clinical efficacy data of multiple therapies with different MoA^24^.

In this study, we develop a QSP model that describes the relationship between cytokines and AD pathogenesis using clinical efficacy data of nine approved or investigational biologic drugs: dupilumab, lebrikizumab, tralokinumab, secukinumab, fezakinumab, nemolizumab, tezepelumab, GBR 830, and recombinant interferon-gamma (rIFNg). We use the model to reveal the pathophysiological backgrounds of dupilumab poor responders and to identify promising drug targets to treat the dupilumab poor responders.

## 2. METHODS

Our QSP model explicitly describes regulatory links between drugs, biological factors, and an efficacy endpoint using graphical scheme and ordinary differential equations (ODEs). The model development processes consist of 1) selecting drugs and biological factors to be described in the model, 2) formulating causal relationship between the biological factors using ODEs, 3) quantifying drug effects on the biological factors, and 4) tuning model parameters of the ODEs so that the model reproduce reference data, which were obtained from published clinical trials.

### 2.1. Selection of drugs and biological factors

We considered only the drugs that showed a higher (not necessarily statistically significant) efficacy than placebo in a placebo-controlled double-blinded clinical study (Table 1, the process is detailed in Supplementary Information (SI) Section 1, Figure S1) and adopted only the highest dose of each drug. For example, we adopted the highest dose, 300 mg weekly, used in the Ph3 study for dupilumab. It allowed us to assess the maximal effects of the MoA and integrate the clinical data from different MoA, rather than investigating the relationship between pharmacokinetics and pharmacodynamics.

**TABLE 1.** Drugs considered in this study

| Drugs | Targets | Dose regimen<br>(highest dose) | Available efficacy endpoints | #patients in placebo/drug arm (Phase) |
| --- | --- | --- | --- | --- |
| Dupilumab<sup>10</sup><br>(anti-IL-4 receptor subunit $\alpha$ antibody) | IL-4 and IL-13 | 300 mg qw, s.c.<br>+TCS | EASI-75,<br>%improved EASI<br>%improved SCORAD | 264/270<br>(Ph3) |
| Lebrikizumab<sup>42</sup><br>(anti-IL-13 antibody) | IL-13 | 250 mg q2w, s.c.<br>+TCS | EASI-75,<br>%improved EASI | 52/75 (Ph2) |
| Tralokinumab<sup>41</sup><br>(anti-IL-13 antibody) | IL-13 | 300 mg q2w, s.c.<br>+TCS | EASI-75<br>%improved EASI | 126/252<br>(Ph3) |
| Secukinumab<sup>16</sup><br>(anti-IL-17A antibody) | IL-17A | 300 mg qw for 4 weeks,<br>followed by 300 mg q4w | %improved EASI<br>%improved SCORAD | 14/27 (Ph2) |
| Fezakinumab<sup>38</sup><br>(anti-IL-22 antibody) | IL-22 | 600 mg at day 0,<br>followed by 300 mg q2w, i.v. | %improved SCORAD<sup>†</sup> | 20/40 (Ph2) |
| Nemolizumab<sup>37</sup><br>(anti-IL-31 receptor subunit $\alpha$ antibody) | IL-31 | 60 mg q4w, s.c. | EASI-75,<br>%improved EASI | 72/143 (Ph3) |
| Tezepelumab<sup>39</sup><br>(anti-TSLP antibody) | TSLP | 280 mg q2w, s.c.<br>+TCS | EASI-75,<br>%improved EASI<br>%improved SCORAD | 56/55 (Ph2) |
| GBR 830<sup>40</sup><br>(anti-OX40L antibody) | OX40L | 10 mg/kg q4w, i.v. | %improved EASI | 16/46 (Ph2) |
| rIFNγ<sup>43</sup> | IFNγ | 50 ug/m2 qd, s.c.<br>+TCS | % improved AD scores<sup>‡</sup> | 43/40 (Ph2) |

†: %improved EASI was estimated from %improved SCORAD using a regression curve (FIGURE S2), ‡: the mean value of the %improved AD scores were regarded as %improved EASI because the evaluated signs were same as those for EASI (erythema, induration, excoriations, and lichenification). When there were multiple clinical studies per drug, we adopted the clinical study of combination therapy with topical corticosteroids, which is more reflective of the likely clinical use compared with monotherapy, and studies with the largest number of patients.
Abbreviations: qw, every week; q2w, every 2 weeks; q4w, every 4 weeks; qd, every day; s.c., subcutaneous administration; i.v., intravenous administration; +TCS, concomitant use of topical corticosteroids.

We did not consider small molecules because they can affect many cytokines, making it difficult to associate clinical efficacy with a specific cytokine in our model. For example, a Janus kinase (JAK) inhibitor, abrocitinib, was excluded in this study, as JAK inhibitors block signaling of a considerable number of cytokines and growth factors.

As biological factors, our model described skin barrier integrity and infiltrated pathogens, which were described as key factors in our previously published mathematical model of AD pathogenesis^26^ as well as the cytokines and OX40L in the skin that have been specifically targeted by the drugs (TABLE 1). Our model also described their related subtypes of Th cells to explicitly represent feedforward and feedback mechanisms (e.g., Th2 secretes IL-4, which promotes T cell differentiation toward Th2).

Some biological factors such as dendritic cells and AMPs were not described as model variables but were taken into consideration as a rationale for regulatory processes in our model (e.g., IL-17A decreases infiltrated pathogens via increasing AMPs), to make the model simpler, yet interpretable. Our model excluded the targets of the excluded drugs because the contribution of those targets on AD pathogenesis has not been clinically confirmed (e.g., IgE was not considered because anti-IgE antibody omalizumab was excluded due to lack of efficacy in clinical trials).

The EASI score was adopted as a model variable to represent an efficacy endpoint.

### 2.2. Formulating causal relationship between biological factors

We developed a mathematical model consisting of 14 ODEs with 51 parameters to simulate the efficacy of the nine drugs (detailed in SI Section 3). The functional relationships between biological factors in the model were described according to published experimental evidence based on human data. We assumed that each cytokine independently affects skin barrier integrity and infiltrated pathogens (e.g., the level of IL-4 does not affect the influence of IL-13 on skin barrier integrity.) as cytokines has shown additive influence on filaggrin expression, which is related to skin barrier integrity and antimicrobial peptides (AMPs), which decrease microbial pathogens^27, 28^.

The model was implemented in Python 3.7.6 (Python Software Foundation).

### 2.3. Modelling drug effects

All the drugs, except for rIFNg, inhibit signaling from biological factors by blocking their binding to the receptors by targeting the cytokine itself or its receptor. Effective concentration of the target biological factor in the skin at time *t, c*(*t*), was modelled by

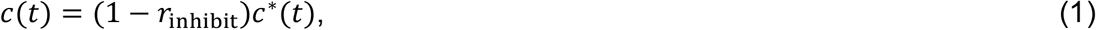

where *c*^∗^(*t*) is the actual concentration of the target biological factor in the skin at *t* and *r*_inhibit_ is the inhibition rate of the target biological factor in the drug treatment. The value of *r*_inhibit_ was determined using the published data on IC_50_ and the mean concentration of drugs in the skin^29^ that was estimated from their concentration in the serum measured in clinical trials (detailed in SI Section 3.4 and TABLE S3). The estimated value of *r*_inhibit_ was 0.99 for all the antibodies, except for *r*_inhibit_=0.44 for tralokinumab (detailed in SI section 3.4).

Administered rIFNg increases the effective amount of IFNg. Effective concentration of IFNg in the skin at *t, c*_IFNg_(*t*), is modelled by

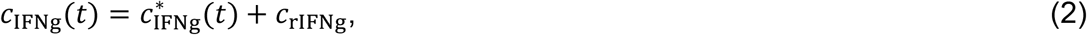

where 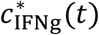 is the actual concentration of IFNg in the skin at *t* and *c*_rIFNg_ is the mean concentration of rIFNg in the skin after rIFNg administration. *c*_rIFNg_ was estimated as 210 based on the pharmacokinetics data of rIFNg^30^ (detailed in SI Section 3.4).

### 2.4. Modelling virtual patients and parameter tuning

We represented each virtual patient by a set of values for the 51 model parameters, where each parameter value is taken from log-normal distribution^31^ (TABLE S4). The probability distribution function, *f*(*k*_*n*_), for the *n*-th parameter, *k*_*n*_, is defined by

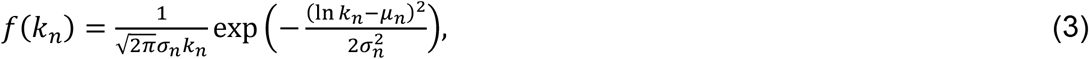

where *μ*_*n*_ and *σ*_*n*_ are the distribution parameters that represent the mean and the standard deviation of ln *k*_*n*_, respectively.

We tuned 102 parameters (*μ*_*n*_ and *σ*_*n*_) that define distributions of the 51 model parameters (detailed in SI Section 4). The 11 parameters, *μ*_*n*_ for elimination rates of the 11 biological factors, were determined using the half-lives measured *in vivo* (serum) in humans (TABLE S5). The remaining 91 parameters were tuned so that the model reproduces the following clinical data;

– The mean and the coefficient of variation of levels of biological factors in observational studies (TABLE S2) and
– The mean EASI scores and EASI-75 when the nine drugs were applied in clinical trials (FIGURE 1).

**FIGURE 1.**
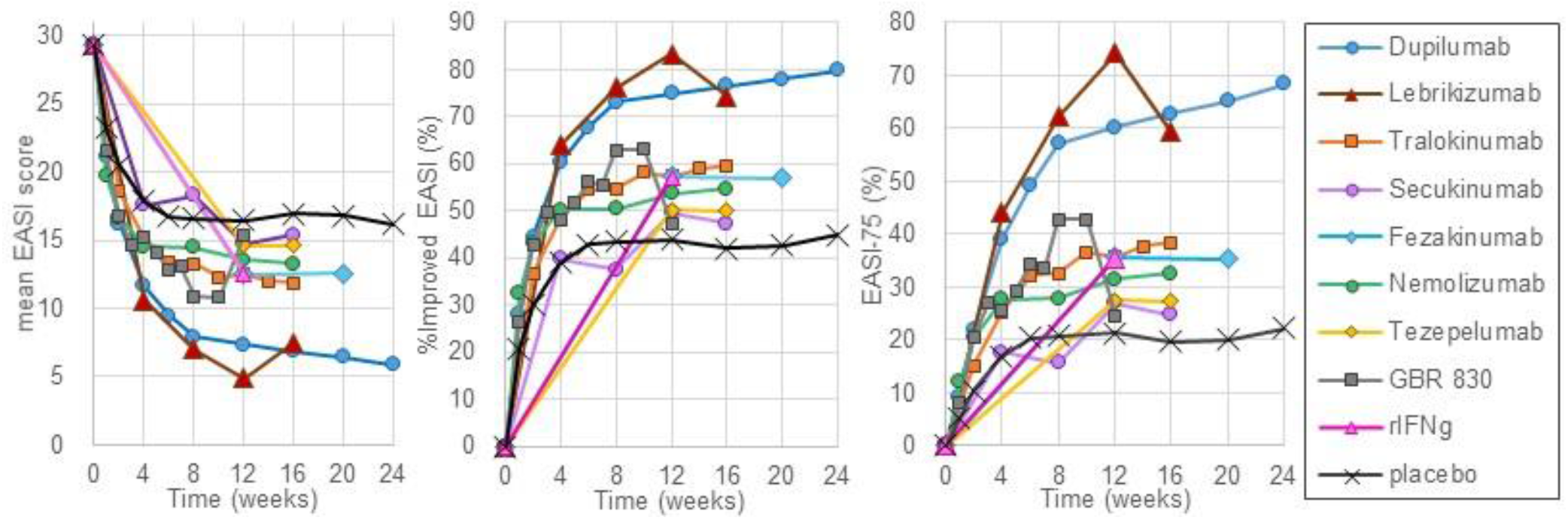
Reference data collected from published clinical trials. The mean EASI score, %improved EASI, and EASI-75 of nine drugs (dupilumab, lebrikizumab, tralokinumab, GBR 830, fezakinumab, rIFNg, tezepelumab, nemolizumab, secukinumab) were normalized according to the method detailed in SI Section 2.

We collected reference data for efficacy endpoints from published clinical trials of the selected drugs. We used EASI-75 (and its related %improved EASI and mean EASI score) as an efficacy endpoint that was adopted as one of the most common primary endpoints in Ph3 studies^10,25^. EASI-75 was normalized to compare clinical efficacies of different clinical trials (detailed in SI Section 2).

Reference data for the levels of biological factors, including cytokines, OX40 ligand (OX40L), and T cells in AD skin lesions, were obtained from human skin biopsy data in published observational studies (TABLE S2). We described protein levels of cytokines and OX40L and count levels of T cells in AD lesion skin by fold change relative to those for healthy subjects or to non-lesional skin of the same AD patients. The skin barrier integrity and infiltrated pathogens were regarded as latent state variables, which have no reference data to be compared with simulated values.

Simulated data for the baseline levels of the biological factors were obtained by simulating steady-state levels of biological factors (at 1000 weeks without drug treatment) using a large number of virtual patients (1000 virtual patients) by randomly sampling each of the parameter values from the distribution in Eq. (3).

Simulated data for the mean EASI and the EASI-75 were obtained by simulating drug treatment for the same number of virtual patients as for the corresponding clinical trial (TABLE 1), while the simulated clinical trial was repeated 1000 times to calculate the 95% confidence interval (CI).

### 2.5. Identification of pathophysiological backgrounds that influence %improved EASI of each drug

To identify the pathophysiological backgrounds of virtual patients that influence %improved EASI of each drug most, we conducted a global sensitivity analysis of the model parameters with respect to %improved EASI. We produced 1000 virtual patients by varying the 51 parameters that represent their pathophysiological backgrounds using Latin hypercube sampling (LHS) and computed partial rank correlation coefficient (PRCC)^32^ between each parameter and %improved EASI of each drug. LHS is a sampling method to explore the entire space of multidimensional parameters efficiently, and PRCC represents a rank correlation coefficient that is controlled for confounding effects that could lead to detecting pseudo-correlations. The evaluated ranges of ln *k*_*n*_ were [*μ*_*n*_ − *σ*_*n*_,*μ*_*n*_ + *σ*_*n*_]. The *p*-values for the PRCC were adjusted for multiple testing with the Bonferroni procedure, where a significance level of adjusted *p* < 0.05 was used.

### 2.6. Simulation of clinical efficacy of hypothetical therapy for dupilumab poor responders

We simulated EASI-75 of a hypothetical therapy for virtual dupilumab poor responders. Virtual dupilumab poor responders were the virtual patients who did not achieve the criterion of EASI-75 (more than 75% improvement of EASI score from week 0) at 24 weeks after dosing dupilumab in 1000 virtual patients. The virtual poor responders were treated with a single drug, combinations of two drugs, and a hypothetical therapy that inhibited combinations of two cytokines or all the cytokines considered in the model (inhibiting IL-4, IL-13, IL-17A, IL-22, IL-31, IFNg, and thymic stromal lymphopoietin: TSLP by 99%).

## 3. RESULTS

### 3.1. Normalization of EASI enabled comparison of clinical efficacy of nine drugs

We selected nine biologic drugs with different MoA that have shown clinical efficacy compared to placebo at least at the population average (TABLE 1) and compared the clinical efficacy of the drugs by mean EASI score, %improved EASI, and EASI-75 after normalization (FIGURE 1). Dupilumab and lebrikizumab showed the highest efficacy among the nine drugs, suggesting that their common target, IL-13, has the highest contribution on AD pathogenesis among the drug targets evaluated in this study. All other drugs also achieved a certain efficacy compared to placebo, confirming the clinical relevance of all the targets to AD pathogenesis.

### 3.2. QSP model reproduced clinical efficacy of nine drugs

We developed a QSP model (FIGURE 2) that describes the MoA for the nine biologic drugs, i.e., regulatory mechanisms between the biological factors and drugs using the published efficacy data from clinical trials.

**FIGURE 2.**
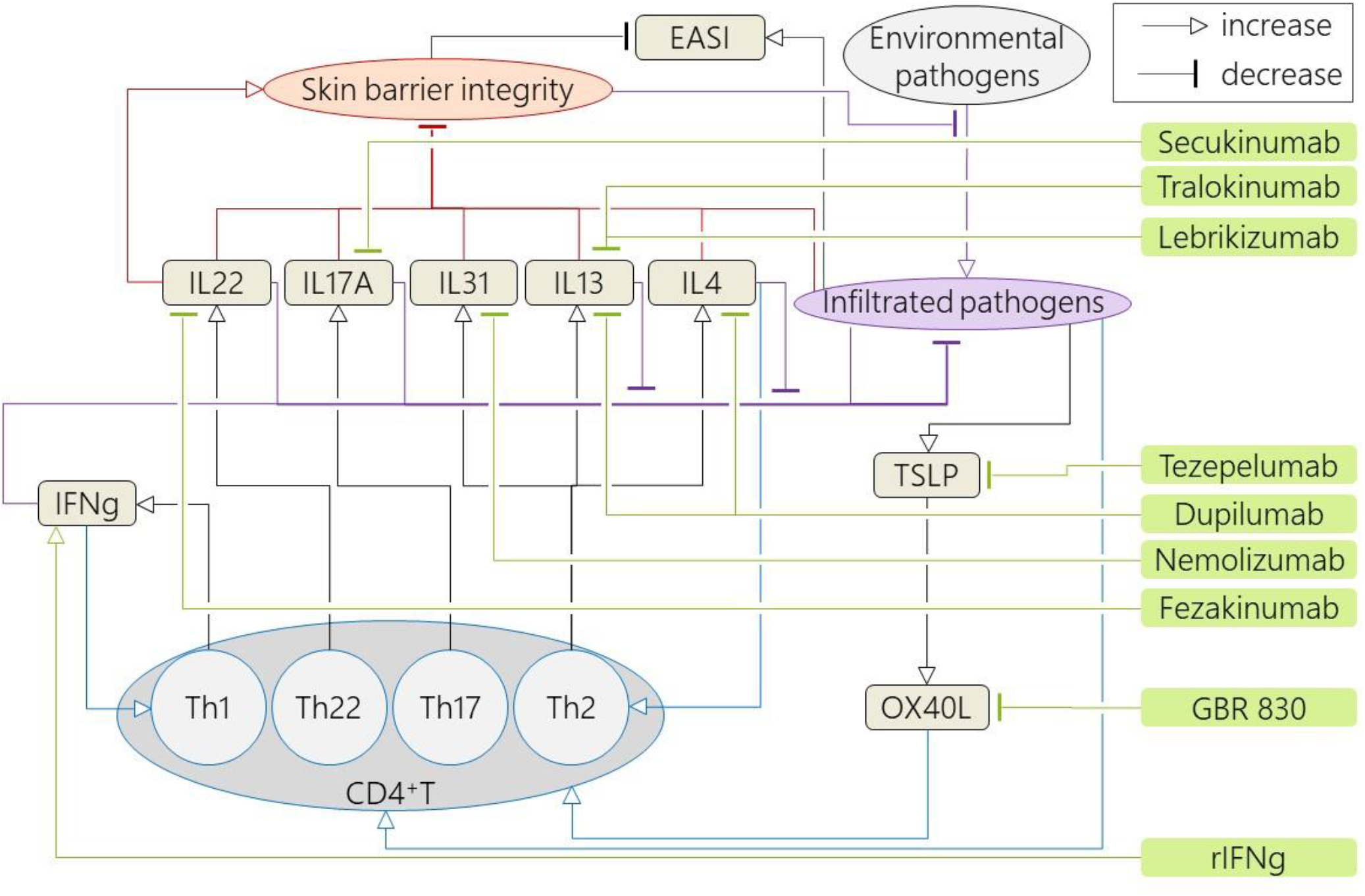
Overview of the proposed mathematical model. The model comprises of an efficacy endpoint (EASI score), biological factors (skin barrier integrity, infiltrated pathogens, cytokines, T cells) and drugs (dupilumab, lebrikizumab, tralokinumab, GBR 830, fezakinumab, rIFNg, tezepelumab, nemolizumab, secukinumab). The functional relationships between biological factors were described according to published human data (details in SI Section 3).

Our model includes 15 biological factors that are targeted by the nine drugs or that are known to be related to AD pathogenesis (TABLE S2). They are seven cytokines (IL-4, IL-13, IL-17A, IL-22, IL-31, IFNg, and TSLP), OX40L, and four subsets of helper T cell (Th1, Th2, Th17, and Th22) that are the main source of the cytokines, except for TSLP which is secreted from keratinocytes. The model also includes “infiltrated pathogens” and “skin barrier integrity” as the main variables for a model of AD pathogenesis^26^, and the EASI score as an efficacy endpoint of each virtual patient. The effects of the drugs were modelled by decreasing or increasing effective concentrations of their target cytokines or OX40L (TABLE S2).

The model contains 51 parameters (e.g., the recovery rate of skin barrier via skin turnover, *k*_1_). We assumed the parameter values, corresponding to the strengths of the regulatory processes, vary between AD patients, so that virtual patients are defined by sets of 51 parameter values. We tuned the distributions of the 51 parameters (TABLE S4) to reproduce the mean and the coefficient of variation of the biological factors in observational studies and mean EASI score and EASI-75 of the drugs (FIGURE 3). The root mean square errors of the mean EASI and EASI-75 between the simulated and reference data were 2.1 (out of 72 = the max EASI) and 7.4%, respectively.

**FIGURE 3.**
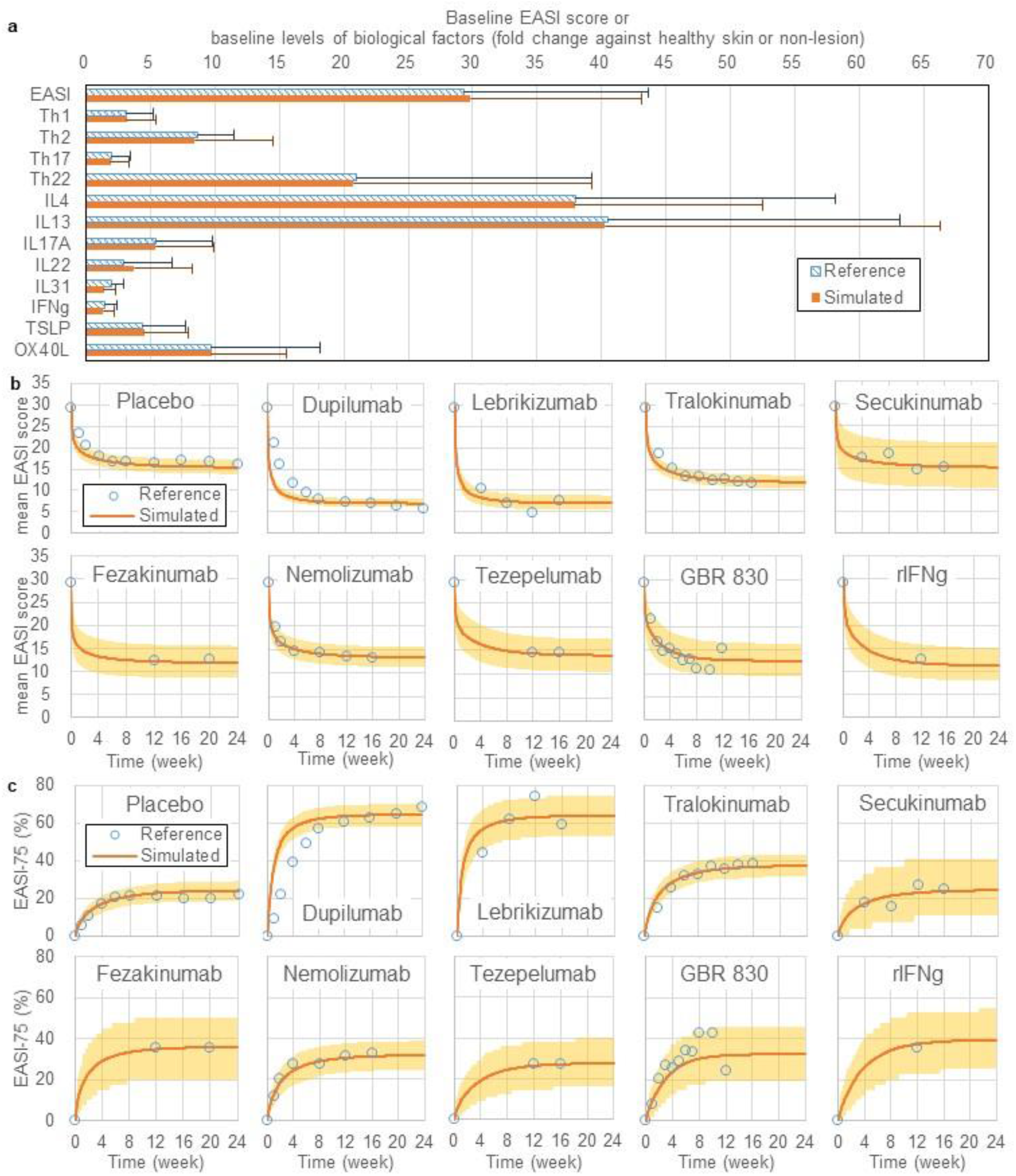
Simulated and reference data of (a) baseline levels of biological factors, (b) mean EASI score, and (c) EASI-75. The model parameters were tuned to minimize the difference between simulated and reference data (detailed in SI section 4.2). (a) Reference data (striped bars) are the measured values of the biological factors in observational studies. Simulated data (filled bars) were obtained by simulating steady-state levels of biological factors (at 1000 weeks without drug treatment) using 1000 virtual patients. Error bars are standard deviations. (b and c) Reference data (unfilled circles) are mean EASI scores (b) and EASI-75 (c) after treatment of each drug. Simulated data for the mean EASI and the EASI-75 were obtained by simulating the efficacies of drug treatment using the same number of virtual patients as for the clinical trials (TABLE 1), and the simulated clinical trial was repeated 1000 times to calculate 95% CI (lines: mean, shaded area: 95% CI).

### 3.3. Global sensitivity analysis detected correlation between turnover rates of IL-13 in the skin and clinical efficacy of dupilumab

The variation in the responsiveness to each drug among the patients is considered to be due to the heterogeneity in the pathophysiological backgrounds of the patients. The responder rates could be improved by patient stratification based on biomarkers that reflect pathophysiological backgrounds^33^. Using the QSP model, we can investigate which pathophysiological backgrounds have influence on clinical efficacy of each drug and which cytokines in the skin can be promising biomarkers for patient stratification. We performed a global sensitivity analysis of the model to investigate the influence of the 51 model parameters (that represent pathophysiological backgrounds of the virtual patients) on %improve EASI of each drug using the LHS-PRCC (FIGURE 4).

**FIGURE 4.**
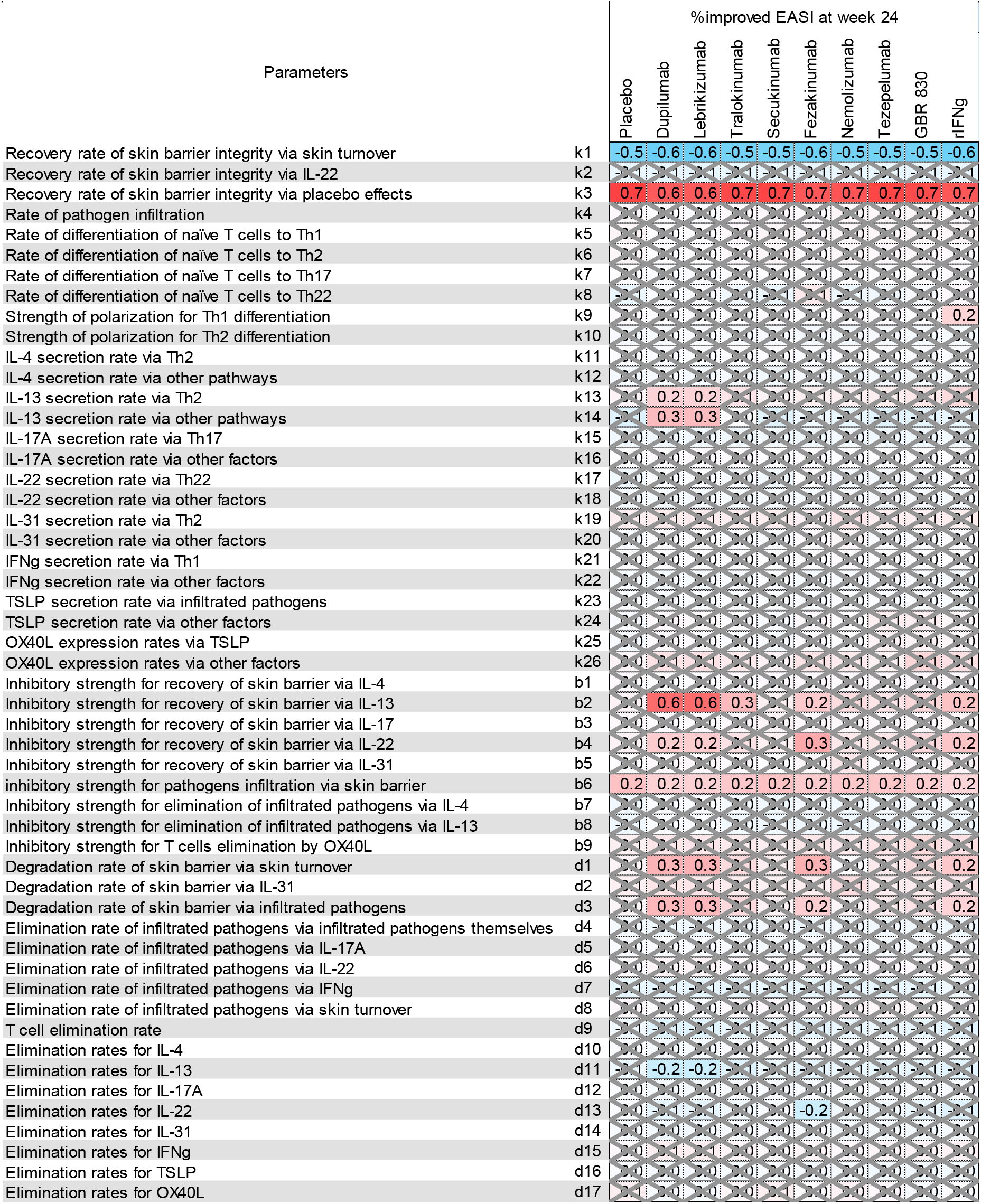
Partial rank correlation coefficient (PRCC) between model parameters and %improved EASI by each drug treatment. 1000 virtual patients (1000 sets of 52 model parameter values) were generated by sampling 52 parameter values independently according to Latin hypercube sampling, and were treated by each drug to simulate %improved EASI at 24 weeks. Open and crossed cells are statistically significant and not significant PRCC (absolute value >0.1 with adjusted *p-*values <0.05), respectively. Positive PRCC means that virtual patients with a higher value of the parameter achieve a higher %improve EASI by the drug treatment (e.g., *k*3). Negative PRCC means that virtual patients with a lower value of the parameter achieve a higher %improve EASI by the drug treatment (e.g., *k*1).

Ten model parameters had a significant PRCC with the %improved EASI by dupilumab (FIGURE 4). Four out of the ten parameters are IL-13-related (*k*_13_, *k*_14_, *b*_2_, and *d*_11_), and the remaining six parameters are skin barrier-related parameters (*k*_1_, *k*_3_, *b*_4_, *b*_6_, *d*_1_, and *d*_3_) that correspond to placebo effects and baseline severity of skin barrier defects rather than each MoA (SI Section 5). The four IL-13-related parameters (*k*_13_, *k*_14_, *b*_2_, and *d*_11_) can characterize responders for dupilumab, as virtual patients with higher *k*_13_, *k*_14_, and *b*_2_ and a lower *d*_11_ were more responsive to treatment by dupilumab. The parameter, *b*_2_, describes the influence of IL-13 on skin barrier damage. IL-4-related parameters (*k*_11_, *k*_12_, *b*_1_, *b*_7_, and *d*_10_) did not have a significant PRCC with %improved EASI by dupilumab, which inhibits both IL-4 and IL-13 signaling, in consistent with the report that clinical efficacy of dupilumab were not correlated with the baseline level of IL-4 mRNA expression^14^. We confirmed that the ten model parameters had different distributions for good and poor responders for dupilumab (SI Section 6).

As three out of the four IL-13 related parameters, *k*_13_, *k*_14_, and *d*_11_, affect the IL-13 baseline level, we hypothesized that baseline levels of some cytokines in the skin could be used as predictive markers of good/poor responders for dupilumab and other drugs. To test this hypothesis, we stratified virtual patients based on their baseline cytokine levels for varying pre-defined threshold values. For the virtual patients whose baseline cytokine level is greater than the threshold value, we simulated the %improved EASI at week 24 of each drug (FIGURE 5). For dupilumab, EASI-75 was improved for patients with a higher IL-13 baseline level. It is consistent with the results from actual clinical trials of dupilumab, where a higher efficacy was observed in the AD patients with higher baseline messenger RNA (mRNA) levels of IL-13^14^.

**FIGURE 5.**
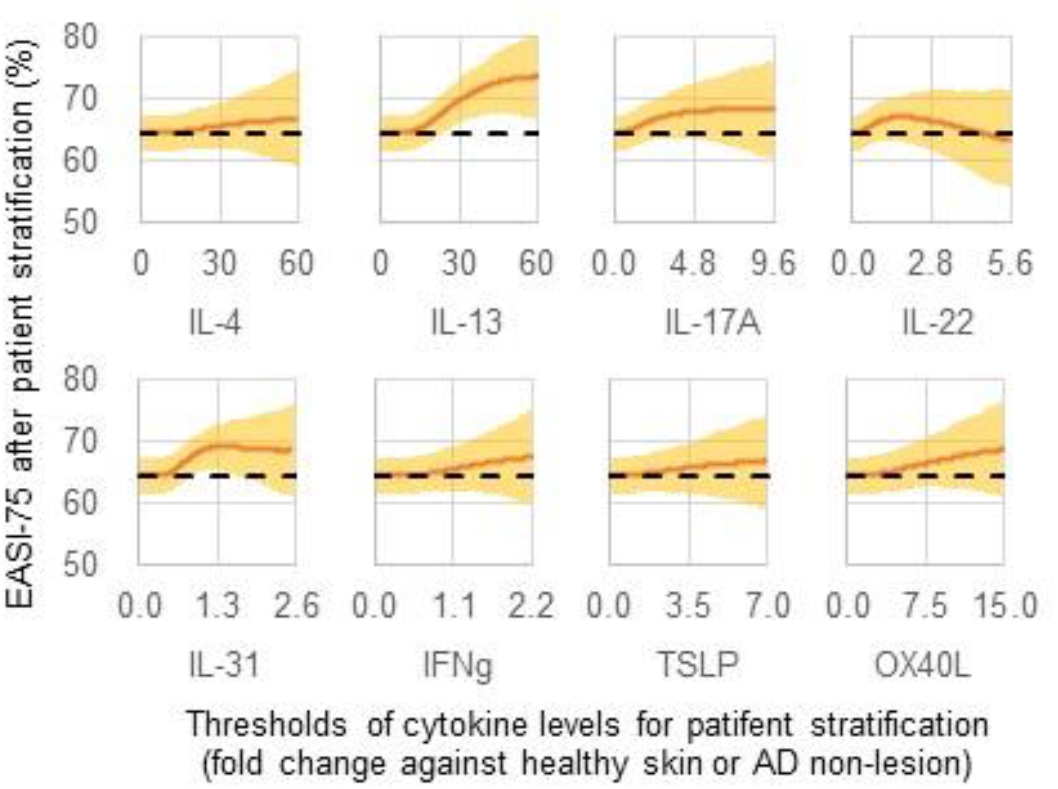
Simulated EASI-75 in dupilumab treatment after patient stratification with different thresholds of cytokine baseline levels. 1000 virtual patients (1000 sets of 52 model parameter values) were generated by randomly sampling each of the parameter values from the distribution in Eq. (3), and were stratified into those whose cytokine baseline level (before dupilumab treatment) was greater than the threshold (x-axis). EASI-75 at 24 weeks (y-axis) were calculated for the stratified patients. The number of stratified virtual patients decreases with an increasing threshold, while the threshold of zero includes all the virtual patients. The maximal threshold for each cytokine was set to ensure that at least 10% of the generated 1000 virtual patients are stratified. Orange solid lines and shaded areas are the mean value and 95% CI of 1000 simulations, respectively. Higher EASI-75 achieved with patient stratification (compared to without stratification with the threshold zero: dashed line) suggests a success in stratifying good responders.

### 3.4. Simulated hypothetical therapies for dupilumab poor responders revealed simultaneous inhibition of IL-13 and IL-22 were effective whereas the nine biologic drugs were ineffective

The proportion of poor responders to dupilumab; the actual and simulated percentages of patients who did not achieve EASI-75 at 24 weeks, was 31%^10^ and 35% [95%CI 30.0%-41.9%. respectively (FIGURE 3). However, therapeutic options for the dupilumab poor responders are limited to increasing topical corticosteroids and adding systemic immunosuppressive agents, although the dupilumab poor responders are often resistant to these treatments and require monitoring for adverse effects^34^.

We hypothesized that the dupilumab poor responders could be responsive to other targeted biologic drugs with different MoA, considering the heterogeneity of AD pathogenesis, and evaluated the potential efficacy of all nine drugs (FIGURE 6a). Every one of the nine drugs failed to show clinical responses in virtual dupilumab poor responders where maximal calculated responder rates, based on EASI-75 at 24 weeks, were only 1.9% [95%CI 0.6%-3.4%. in fezakinumab. These low efficacies imply an upper efficacy ceiling for drugs that target single cytokines to treat dupilumab poor responders.

**FIGURE 6.**
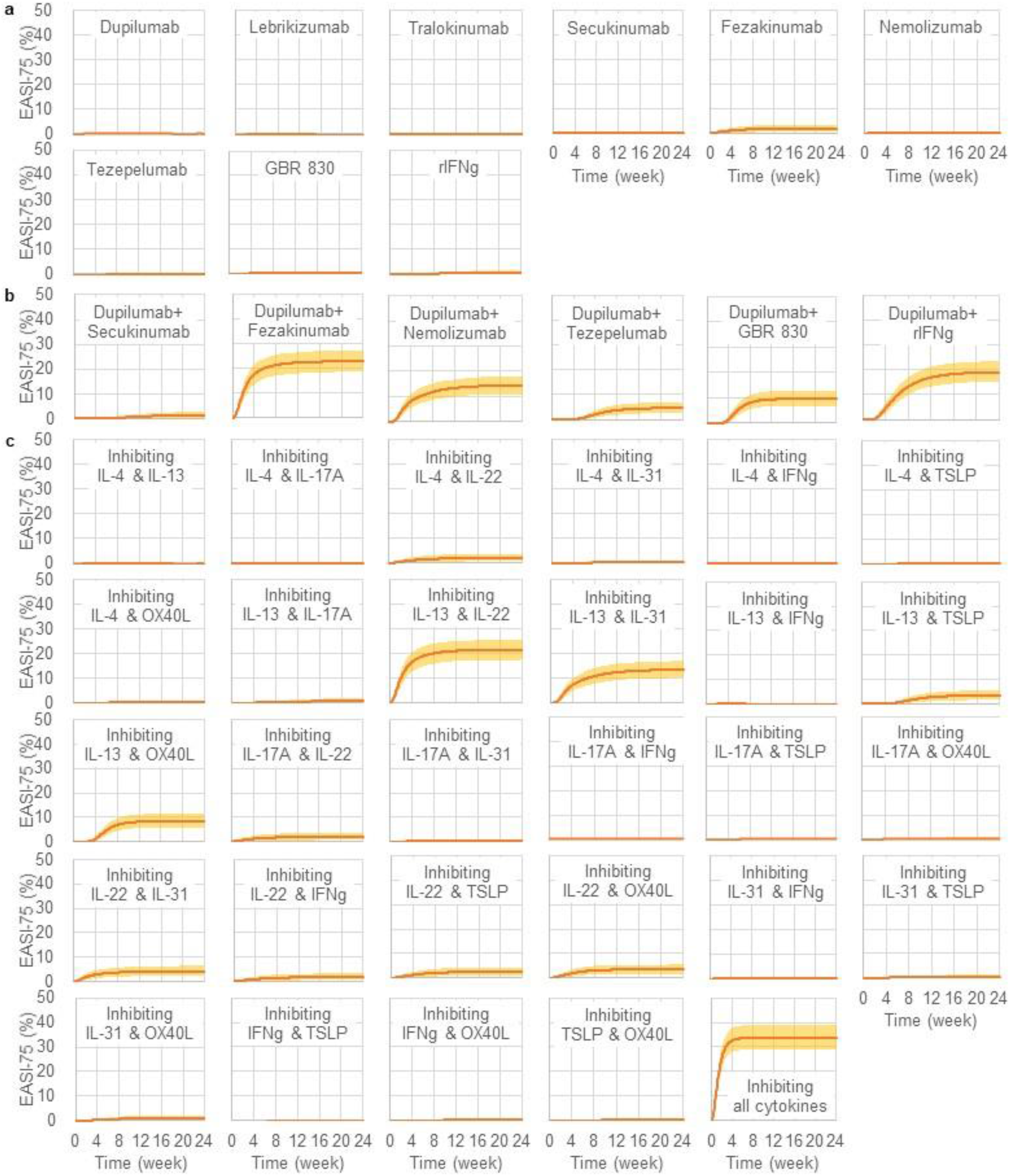
Simulated EASI-75 in virtual dupilumab poor responders. The virtual dupilumab poor responders were treated in simulated clinical trials with (a) a single drug, (b) a combination of two drugs, and (c) blocking a combination of two cytokines or all the cytokines (inhibiting IL-4, IL-13, IL-17A, IL-22, IL-31, IFNg, and TSLP by 99%). Virtual dupilumab poor responders were the virtual patients who did not achieve the EASI-75 criterion at 24 weeks among 1000 virtual patients (mean 356 with 95% CI 300-419 virtual patients). The simulated clinical trial was repeated 1000 times to obtain 95% CI. Lines and shaded areas are the mean values and 95% CI of 1000 simulations, respectively.

We then evaluated the potential efficacy of hypothetical therapies that inhibit two cytokines, mimicking a bispecific antibody, for the virtual dupilumab poor responders. Our simulation demonstrated that dosing one additional drug to dupilumab achieved a better efficacy for virtual dupilumab poor responders (FIGURE 6b). The maximal EASI-75 in the virtual dupilumab poor responders (0% achieving EASI-75 at 24 weeks with dupilumab therapy) at 24 weeks was 23.0% [95%CI 18.4%-27.3%. in dupilumab + fezakinumab, implying that inhibition of both IL-13 and IL-22 would be a promising combination for treatment of dupilumab poor responders. Indeed, inhibition of both IL-13 and IL-22 showed the highest clinical responses among all the combinations of two cytokines (FIGURE 6c), with EASI-75 at 24 weeks being 21.6% [95%CI 17.4%-25.5%.. These results are in concordance with a clinically observed negative correlation between the clinical efficacy of dupilumab and the baseline level of IL-22 mRNA expression (not significant, rank correlation coefficient −0.208 with *p*-value of 0.422) ^14^. The simulation results, in combination with the clinical observation^14^, suggest that inhibition of IL-22 in addition to the dupilumab treatment is effective for dupilumab poor responders.

We also confirmed that blocking all targeted cytokines (inhibiting IL-4, IL-13, IL-17A, IL-22, IL-31, IFNg, and TSLP by 99%) achieved a higher EASI-75 (33.8% [95%CI 28.8%-38.7%., FIGURE 6c). The responder rate did not reach 100% even when all the cytokines were blocked because some cytokines have not only detrimental but also beneficial effects. For example, IL-17A and IL-22 damage the skin barrier by inhibiting epidermal differentiation (detrimental effects) while they decrease infiltrated pathogens via increasing AMP (beneficial effects). Hence, inhibition of cytokines can partly exacerbate the AD symptoms, suggesting that finding optimal combinations of drugs requires a systems-level investigation.

## 4. DISCUSSION

### 4.1. QSP model to simulate biologics efficacy

Several biologic drugs targeting AD pathogenic cytokines have been developed and shown clinical efficacy to some extent in AD patients. Although only one biologic drug, dupilumab, has been currently approved, it is useful to conduct model-based meta-analysis by integrating the results from clinical trials of other biologic drugs, including those that failed to show clinically significant efficacy and those under development, to enhance the understanding of AD pathogenesis and clinical efficacy of drugs with different MoA. In this study, we conducted model-based meta-analysis of clinical trials on the nine biologic drugs for AD (dupilumab, lebrikizumab, tralokinumab, secukinumab, fezakinumab, nemolizumab, and tezepelumab, GBR830, and rIFNg) that were published by Dec 2020 (FIGURE S1), and developed a QSP model of biologics efficacy in AD patients (FIGURE 2). This QSP model describes dynamic relationships between biological factors and clinical efficacies by integrating knowledge obtained from experiments using human samples and clinical trials of multiple drugs. The model reproduced the clinical efficacies of the biologic drugs observed in clinical trials and baseline levels of biological factors (e.g., cytokines) in the skin from published studies (FIGURE 3).

### 4.2. Comparison of clinical efficacies of biologics

Comparison of clinical efficacies (FIGURE 1) demonstrated that dupilumab and lebrikizumab showed the highest efficacy among the nine drugs investigated in this study, suggesting that their common target, IL-13, is the most crucial target to improve AD severity scores. The comparable efficacy between dupilumab (inhibiting both IL-4 and IL-13) and lebrikizumab (inhibiting IL-13 only) may suggest inhibition of IL-4 signaling has a minor contribution on the efficacy of dupilumab. Another anti-IL-13 antibody, tralokinumab, showed a significantly lower clinical efficacy than lebrikizumab. Lebrikizumab inhibits binding of IL-13 to IL-13Rα1 only, while tralokinumab inhibits binding to both IL-13Rα1 and IL-13Rα2. IL-13Rα1 forms a heterodimeric receptor with IL-4Rα and is related to the effects of IL-13 signaling in AD pathogenesis while IL-13Rα2 is a decoy receptor to decrease IL-13 signaling via IL-13Rα1. Hence, tralokinumab not only inhibits IL-13 signaling via IL-13Rα1 but also enhances IL-13 signaling via inhibition of IL-13 binding to IL-13Rα2^35^. In our simulation, effective inhibition of IL-13 signaling by tralokinumab was estimated 44% of lebrikizumab (*e*_a2_ = 0.44). The difference in the efficacy between lebrikizumab and tralokinumab may come from different mechanisms for IL-13 inhibition, dosing regimens, or trial design.

### 4.3. Pathophysiological backgrounds of dupilumab poor responders

This study presents a QSP-based approach to identify potential predictive biomarkers of clinical efficacy through mechanistic model-based simulation. We used the developed model to explore pathophysiological backgrounds of dupilumab poor responders. Our simulation demonstrated that the higher responder rates for dupilumab are expected in patients with the higher baseline level of IL-13 in the skin (FIGURES 4 and 5). Although it also showed baseline levels of other cytokines (e.g., IFNg) in the skin influence responder rates, these influences may be due to a pseudo-correlation of the cytokines with clinical efficacy because the parameters related to those cytokines were not detected as important in the global sensitivity analysis, which evaluates the “causative” influence of the parameters on efficacy. The cytokines detected due to the pseudo-correlation should not be used as predictive biomarkers for patient stratification, because such pseudo-correlations may have low reproducibility. Hence, our simulations suggest that the baseline level of IL-13 in the skin would be more suitable than other cytokines to be used as a biomarker for patient stratification in dupilumab treatment.

### 4.4. Simulated efficacy of hypothetical therapy in dupilumab poor responders

We also used the model to investigate alternative therapeutic options for virtual dupilumab poor responders. Our simulation results suggested inhibition of a single cytokine would be insufficient to achieve clinical response in virtual dupilumab poor responders, whereas inhibition of multiple cytokines could achieve clinical response (FIGURE 6a,b). Especially, IL-13 and IL-22 were identified to be the best combination to treat virtual dupilumab poor responders (FIGURE 6c). Inhibition of two cytokines can be realized by a bispecific antibody as well as combination of two antibodies.

Our simulated hypothetical therapy that inhibits all the cytokines in this model (IL-4, IL-13, IL-17A, IL-22, IL-31, IFNg, and TSLP) showed higher clinical efficacy than inhibiting single and a combination of two cytokines (FIGURE 6c). These results suggest that inhibiting multiple cytokines can exert higher efficacy than biologic drugs targeting only one or two cytokines. In fact, the approach to inhibit signaling of multiple cytokines has been already realized by JAK inhibitors. For instance, abrocitinib blocks JAK1, which is an intracellular tyrosine kinase linked to intracellular domains of many cytokine receptors, including IL-4, IL-13, IL-22, IL-31, IFNg, and TSLP. Abrocitinib showed comparable efficacy with dupilumab^11^, but there were still a significant percentage of poor responders. One of the reasons why even multiple cytokine inhibition cannot achieve clinical response in all AD patients is that inhibition of cytokines can deteriorate AD symptoms because some cytokines have both detrimental and beneficial effects in AD pathogenesis and the contributions of each cytokine on AD pathogenesis would vary among the AD patients. Hence, appropriate drugs would vary according to the pathophysiological backgrounds of the patients. Patient stratification may be beneficial not only for biologic drugs but also for the drugs that target multiple cytokines.

### 4.5. Limitation of this study using the model

We used clinical efficacy results of the biologic drugs as the reference data to tune the model parameters. We adopted Ph2 as well as Ph3 studies to make a maximal use of available clinical data (TABLE 1). However, the results from Ph2 studies with a rather small number of patients need to be confirmed with those from Ph3 studies with a larger number of patients. Our model assumptions include that virtual patients were generated from single modal distributions of the model parameters. There could be a multimodal distribution due to the genetic backgrounds or other demographic variances in a real population of AD patients. For the practical purposes of generating this model we assumed the %improved EASI was comparable across clinical trials. We know that outcome measures may be influenced by the concomitant use of topical corticosteroids and the statistical methods used to adjust for or censor topical corticosteroid use and to impute missing data. The model will need to be updated according to availability of the reference data and assumptions if these are changed as new data emerge.

### 4.6. Prospects for model-informed drug development

The proposed model could be further expanded for future development of new drugs for AD, by including new drug targets and pharmacokinetics and pharmacodynamics profiles of new drugs. The expanded model will be a useful tool to computationally evaluate potential clinical efficacies of new drug candidates, to identify biomarkers for patient stratification, to clarify the difference from existing drugs (e.g., showing significant efficacy in dupilumab poor responders), and to design optimal doses with considering variations in pathophysiological backgrounds.

Such simulated data may inform design of future clinical trials. For example, simulated variability in drug efficacy may help determine the number of participants and the study periods required, comparison of the simulated efficacy for new drug candidates and competitive drugs may predict the probability of success in achieving clinical efficacy, and simulated patient stratification may determine pathophysiological backgrounds of patients to be recruited. The model simulation may also enhance understanding of clinical trial results. For example, sensitivity analysis of the model may identify pathophysiological backgrounds that affect the individual variability in pharmacokinetics and clinical efficacy (e.g., this study identified IL-13-related parameters to be related with the dupilumab efficacy). Furthermore, it could inform novel therapeutic approaches as a reverse translational research (e.g., simultaneous inhibition of IL-13 and IL-22 proposed in this study).

The model contributes to a systems-level understanding of AD pathogenesis and the drug effects and to evaluate the effects of new potential drug targets by computational simulation. This could serve as model-informed drug development^36^ for precision medicine. The code of the QSP model is available at https://github.com/Tanaka-Group/AD_QSP_model.

## Data Availability

The code of the QSP model is available.

https://github.com/Tanaka-Group/AD_QSP_model

## Acknowledgments

We thank Dr Elisa Domínguez-Hüttinger for her insightful comments on our manuscript.

