## Supplementary Information for "A mathematical model to identify optimal combinations of drug targets for dupilumab poor responders in atopic dermatitis"

### 1. Selection of clinical studies

We selected clinical studies to be referenced in the QSP model according to pre-defined inclusion and exclusion criteria (FIGURE S1).

We searched the clinical trials that evaluated efficacy of biologic drugs in AD patients, where Ph2 studies were not searched if Ph3 results were available and case reports were not searched if placebo-controlled double-blind clinical trial data were available. We excluded investigational drugs if the clinical efficacies were not evaluated by a placebo-controlled study, if they failed to show a clinical efficacy compared to placebo, if the clinical trials recruited only a small number (<10) of AD patients, if evaluation periods were short (less than 4 weeks), or if data of efficacy endpoints were not published (TABLE S1). When there remained multiple clinical studies per drug, we adopted the clinical study of combination therapy with topical corticosteroids, which is more reflective of the likely clinical use compared with monotherapy, and studies with the largest number of patients.

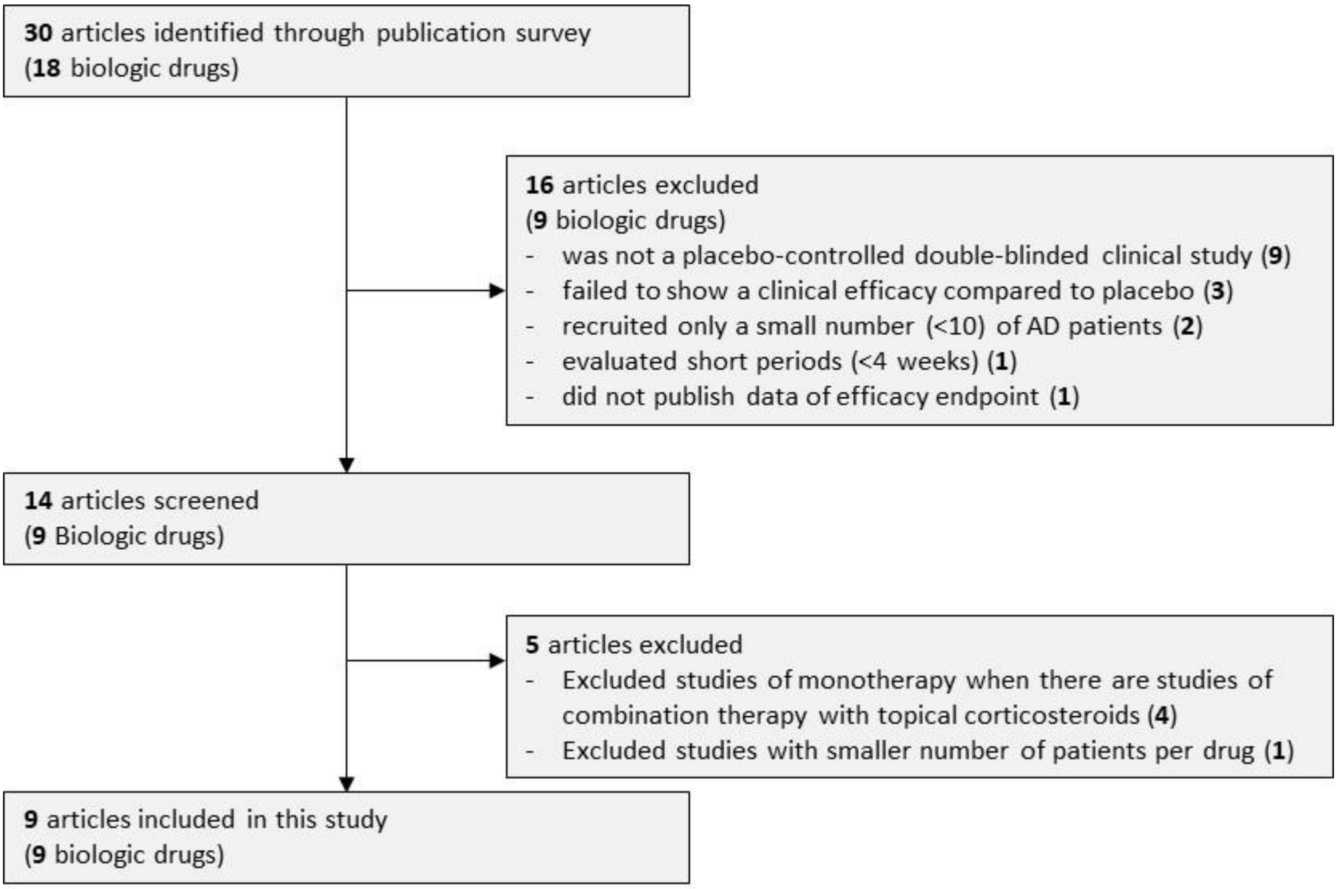


**FIGURE S1** Clinical studies selection process

### **TABLE S1** Drugs excluded in this study

| Drugs | Targets | #patients in placebo/drug arm (Phase) | Reasons to be excluded |
|---|---|---|---|
| Omalizumab [エラー! 参照元が見つかりません。, 2]<br>(anti-IgE antibody) | IgE | 4/4 and 7/13 (Ph4) | • It is difficult to interpret the results due to the small number of patients<br>• Omalizumab showed comparable or lower clinical efficacy than placebo in terms of IGA score and %improved SCORAD (not significant) |
| Mepolizumab<br>(anti-IL5 antibody) | IL-5 | 16/18 at baseline<br>11/11 at week 12<br>8/6 at week 16<br>4/4 at week 20 (Ph2)[3]<br>23/20 (Ph2)[4] | • It is difficult to interpret the results due to the small number of patients after 12 weeks[3]<br>• Evaluation periods were short (2 weeks)[4] |
| MOR106[5]<br>(anti-IL-17C antibody) | IL-17C | N/A (Ph2) | • Data of the primary endpoint are unavailable<br>• The interim analysis detected a low probability to meet the primary endpoint (%improve EASI) |
| Etokimab[6]<br>(anti-IL-33 antibody) | IL-33 | 60/59-61 (Ph2) | Etokimab showed a lower clinical efficacy than placebo in terms of %improve EASI at several dose regimens including the highest dose (not significant) |
| Tocilizumab[7]<br>(anti-IL-6 antibody) | IL-6 | -/3 | • Placebo-controlled double-blind clinical trial data are unavailable.<br>• A single-arm trial of three AD patients showed tocilizumab significantly improved the EASI score compared with baseline but it is difficult to interpret the results due to the small number of patients |
| Ustekinumab<br>(anti-IL-12/23 antibody) | IL-12 and IL-23 | 27/24-28 (Ph2)[8]<br>16/16 (Ph2)[9] | Ustekinumab showed only comparable clinical efficacy to placebo in terms of %improve EASI (placebo: 37.5% vs. Ustekinumab: 38.2%-39.8% at week 12, not significant)[8] and SCORAD (less decreased in ustekinumab compared with placebo on average, not significant)[9] |
| Infliximab<br>(anti-TNFa antibody) | TNFa | -/9[10], 1[11] | Placebo-controlled double-blind clinical trial data are unavailable (A single-arm trial of nine AD patients showed infliximab significantly improved the EASI score compared with baseline) |
| Etanercept | TNFa | -/2[12], 2[13] | Placebo-controlled double-blind clinical trial data are unavailable |
| Rituximab<br>(anti-CD20 antibody) | B cells | -/6[14], 2[15], 3[16] | Placebo-controlled double-blind clinical trial data are unavailable (A single-arm trial of six AD patients showed rituximab significantly improved the EASI score compared with baseline) |

### 2. Data processing

#### 2.1. Conversion of reported efficacy endpoints to %improved EASI

All the drugs, except for fezakinumab and rIFNg, reported %improved EASI as an efficacy endpoint.

For fezakinumab, we estimated %improved EASI from %improved SCORAD using a regression curve obtained from the relationship between %improved EASI and %improved SCORAD in clinical trials of multiple drugs[1, 17, 18, 19] (FIGURE S2).

For rIFNg, we substituted %improved EASI by the mean value of the %improved scores of the disease signs evaluated (erythema, induration, excoriations, and lichenification)[20].

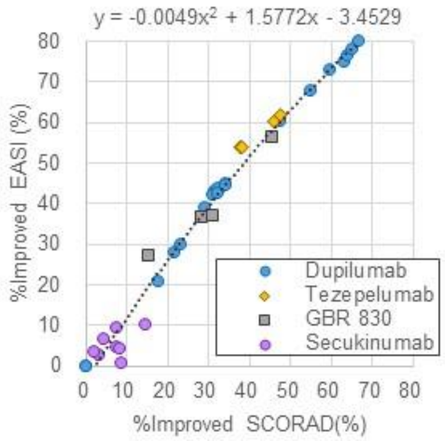


**FIGURE S2** Regression curve to estimate %improved EASI from %improved SCORAD

#### 2.2. Normalization of %improved EASI

%improved EASI was normalized to compare clinical efficacies evaluated in different clinical trials. We assumed that the observed %improved EASI is a sum of %improved EASI from net effects of each drug and that from placebo effects, and described normalized %improved EASI, $e_i(t)$, for the *i*-th drug at time *t* by

$$e_i(t) = (e_i^*(t) - e_{\mathrm{P}_i}^*(t)) + e_{\mathrm{P_d}}^*(t). \qquad \text{(S1)}$$

The first term corresponds to the difference of the efficacy (%improved EASI) between drug ($e_i^*(t)$) and placebo groups ($e_{\mathrm{P}_i}^*(t)$) to adjust for the different extent of the efficacy in the placebo group in each clinical study due to differences in patient background, concomitant drugs, and sites of study[21]. We then added the same extent of the efficacy in the placebo group ( $e_{\mathrm{P_d}}^*(t)$ in the dupilumab clinical trial) in this virtual simulation so that the normalized %improved EASI represents the scale of the observed %improved EASI (the sum of %improved EASI from net effects of each drug and that from placebo effects).

#### 2.3. Normalization of mean EASI score

Normalized mean EASI score was calculated using the baseline mean EASI score (the mean EASI score before the trial) in dupilumab clinical trial[1] and the normalized %improved EASI as follows:

$$a_i(t) = \frac{a_{\mathrm{d}}(0)(100 - e_i(t))}{100} \qquad \text{(S2)}$$

where $a_i(t)$ is the normalized mean EASI score of $i$-th drug at $t$, $a_\mathrm{d}(0)$ is the reported baseline mean EASI score in the dupilumab clinical trial (the mean value of placebo- and dupilumab-treated groups).

#### 2.4. Normalization of EASI-75 using %improved EASI

Normalized EASI-75 was estimated from the normalized %improved EASI using a regression curve obtained from the relationship between %improved EASI and EASI-75 in clinical trials of multiple drugs[1, 22, 18, 19, 23] (FIGURE S3). The relationship between %improved EASI and EASI-75 is affected by the variation of %improved EASI among the patients. Relatively large deviation of Tezepelumab and GBR 830 in the relationship may derive from the smaller variation between the patients due to the small number of patients in the Ph2 studies compared with the Ph3 studies (dupilumab, nemolizumab, and tralokinumab).

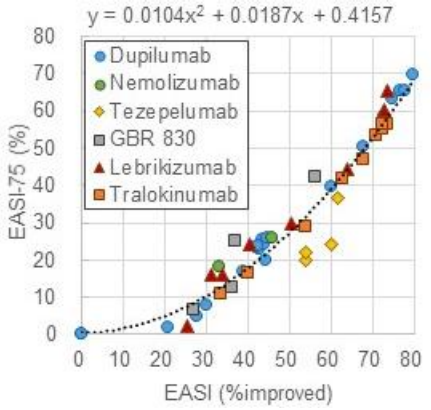


**FIGURE S3** Regression curve to estimate EASI-75 from %improved EASI

### 3. Model structure

The model (FIGURE 2) is formulated by Eqs. (S3)-(S17) shown below. This section details components of the model: EASI score (the output of the model), skin barrier integrity and infiltrated pathogens, cytokines, T cells and drug effects (TABLE S2).

In this model, $t$ is the time after the start of drug treatment. The baseline levels of biological factors ($t$=0) were obtained from the simulated steady-state level (after 1000 weeks) without any intervention. We referred to the reported levels of biological factors in observational studies as the reference values for the baseline levels, assuming that levels of the biological factors were stable before the start of drug treatments. We described protein levels of cytokines and OX40L and count levels of T cells in AD lesion skin by fold changes relative to those for healthy subjects or to non-lesional skin of the same AD patients in order to cancel out the effects of difference measurement methods and units used in different studies, and to avoid using an ambiguous unit of 'T cell count/field of view' in the immunohistochemistry.

**TABLE S2** Biological factors as model variables

| Factors | Model variables | Reason for inclusion | Reported levels in AD lesion | |
|---|---|---|---|---|
| | | | Mean (%CV) | Units |
| IL-4 | $c_4(t)$ | Target of dupilumab | 38.0 (53) [24 a,d] | Fold change against healthy skin |
| IL-13 | $c_{13}(t)$ | Target of dupilumab, tralokinumab, and lebrikizumab | 40.5 (56) [24 a,d] | Fold change against healthy skin |
| IL-17A | $c_{17}(t)$ | Target of secukinumab | 5.4 (81) [24 a,d] | Fold change against healthy skin |
| IL-22 | $c_{22}(t)$ | Target of fezakinumab | 3.0 (124)[25 e,d] | Fold change against healthy skin |
| IL-31 | $c_{31}(t)$ | Target of nemolizumab | 2.0 (49)[25 b,d] | Fold change against healthy skin |
| IFNg | $c_g(t)$ | Target of rIFNg | 1.5 (62)[24 a,d] | Fold change against healthy skin |
| TSLP | $c_{TS}(t)$ | Target of tezepelumab | 4.4 (76)[26 c] | Fold change against healthy skin |
| OX40L | $c_{OX}(t)$ | Target of GBR 830 | 9.7 (87)[27 b] | Fold change against AD non-lesion |
| Th1 cells | $c_{t1}(t)$ | Main source of IFNg | 3.1 (68)[28, 29 b] | Fold change against AD non-lesion |
| Th2 cells | $c_{t2}(t)$ | Main source of IL-4, IL-13, and IL-31 | 8.7 (32)[28, 29 b] | Fold change against AD non-lesion |
| Th17 cells | $c_{t17}(t)$ | Main source of IL-17A | 2.0 (76)[28, 29 b] | Fold change against AD non-lesion |
| Th22 cells | $c_{t22}(t)$ | Main source of IL-22 | 21.0 (87)[28, 29 b] | Fold change against AD non-lesion |
| Infiltrated pathogen | $p(t)$ | Key factor in the previous model<br>Its amount is affected by cytokines via AMP | - [e] | - |
| Skin barrier integrity | $s(t)$ | Key factor in the previous model<br>It protects skin from infiltrating pathogens<br>It is related to severity of AD lesion | - [e] | - |
| EASI score | $e(t)$ | Clinical endpoints | 29.3 (49)[1 b,d,f] | - |

a: mild-to-moderate AD patients. b: moderate-to-severe AD patients. c: both mild-to-moderate and moderate-to-severe AD patients. d: %CV was estimated from IQR. e: no reference data to be compared with simulated values. f: mean baseline value of dupilumab treatment: 29.0 and placebo treatment: 29.6 of AD patients in dupilumab clinical trial

#### 3.1. EASI score, skin barrier integrity, and infiltrated pathogens

We consider EASI as the treatment outcome that will be compared directly to the data from clinical trials.

EASI score (ranging from 0 to 72), is calculated using the severity and the area scores of equally weighted four AD signs (erythema, induration, excoriations, and lichenification) on four body regions (head/neck, trunk, upper limbs, and lower limbs) [30]. In our model, EASI score, $e(t)$, is described (FIGURE S4) by

$$e(t) = 72\,\frac{2p(t)+2(1-s(t))}{4}, \qquad \text{(S3)}$$

where 72 is the maximal EASI score, $p(t)$ is the concentration of infiltrated pathogens with a range from 0 (pathogen-free) to 1 (maximal infiltration) at $t$, and $s(t)$ is the level of skin barrier integrity with a range from 0 (complete destruction) to 1 (healthy state) at $t$. Extents of two AD signs (erythema and induration) and those of remaining two signs (excoriations and lichenification) were surrogated by $p(t)$ and 1- $s(t)$, respectively, as described below. We set $e(0) = 29.3$, the baseline EASI score of the participants in the dupilumab clinical trial, which was used as a reference value to normalize EASI scores in all clinical trials.

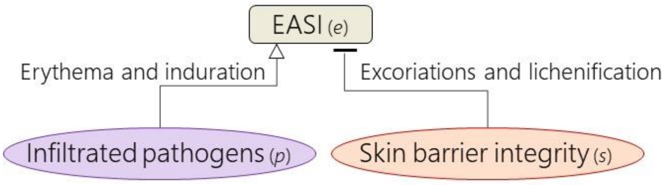


**FIGURE S4** Relationship between EASI score and infiltrated pathogens and skin barrier integrity in our model.

We assumed erythema and induration correlate with the infiltrated pathogen load ($p(t)$) because these two signs can be induced by *Staphylococcus aureus*[31], which is commonly colonized in AD skin lesion[32]. Erythema is caused by inflammatory vasodilation by histamines[33]. Histamine is released mainly from mast cells that are activated by detecting infiltrated pathogens as antigens through antigen-specific IgE[34]. Although both infiltrated pathogens and IgE play a role in this process, we associated the released histamine concentration with infiltrated pathogens load in this model because the amount of histamine release is more dependent on the amount of antigens than that of IgE[35]. Low contribution of IgE on the AD pathogenesis is also suggested by a lack of clinical efficacy demonstrated for omalizumab (IgE neutralizing anti-IgE antibody). The other two signs, excoriations and lichenification are caused by scratching[36], which damages skin barrier integrity. The degree of damage of the skin barrier integrity is described by $1 - s(t)$.

The dynamics of $s(t)$ and $p(t)$ that determine $e(t)$ are described below.

(a) Skin barrier integrity

The dynamics of the skin barrier integrity, $s(t)$, is described (FIGURE S5) by

$$\frac{ds(t)}{dt} = \frac{(1-s(t))(k_1 + k_2 c_{22}(t) + k_3)}{(1+b_1 c_4(t))(1+b_2 c_{13}(t))(1+b_3 c_{17}(t))(1+b_4 c_{22}(t))(1+b_5 c_{31}(t))} - s(t)\{d_1(1 + d_3 p(t)) + d_2 c_{31}(t)\}, \quad \text{(S4)}$$

where $c_4(t)$, $c_{13}(t)$, $c_{17}(t)$, $c_{22}(t)$, and $c_{31}(t)$ are the concentrations of IL-4, IL-13, IL-17A, IL-22, and IL-31, respectively, $k_1$, $k_2$, and $k_3$ are the recovery rates of skin barrier integrity via skin turnover, IL-22, and placebo effects, respectively, $d_1$, $d_2$, and $d_3$ are the degradation rates of skin barrier via skin turnover, IL-31, and infiltrated pathogens, respectively, and $b_1$, $b_2$, $b_3$, $b_4$, and $b_5$ correspond to the inhibitory strengths for recovery of skin barrier via IL-4, IL-13, IL-17A, IL-22, and IL-31, respectively. Levels of skin barrier integrity were defined between 0 (completely damaged) and 1 (healthy).

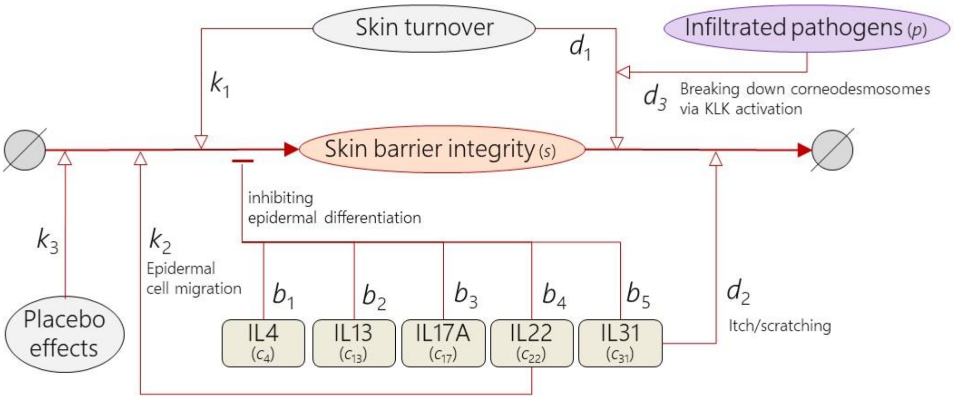


**FIGURE S5** Regulation of skin barrier integrity level

Our model assumes that skin barrier integrity recovers by skin turnover (with the recovery rate, $k_1$), epidermal cell migration ($k_2$), and placebo effects ($k_3$). We assumed that skin turnover occurs independently from external perturbation and epidermal cell migration is promoted by IL-22[37]. The placebo effect was applied to the models for both placebo- and drug-treated groups, as placebo-treated patients demonstrated improvement of the EASI score, presumably because of the controlled care for AD patients with concomitant drugs such as emollients in the clinical trials.

We modelled compromise the recovery of skin barrier integrity by IL-4[38] (with the strength $b_1$), IL-13[39] ($b_2$), IL-17A[40] ($b_3$), IL-22[37] ($b_4$), and IL-31[41] ($b_5$), as they are shown to decrease filaggrin production and inhibit epidermal differentiation.

We also modelled degradation of the skin barrier by skin turnover (with the degradation rate $d_1$) and scratching through itch induced by IL-31 ($d_2$)[42]. Our model assumed that the impairment of skin barrier by skin turnover ($d_1$) is potentiated by infiltrated pathogens ($d_3$), such as *Staphylococcus aureus*, as *Staphylococcus aureus* stimulates TLR2[43] and thereby excessively activates KLK5[44] to increase desquamation in the skin turnover process[45].

(b) Infiltrated pathogens

The dynamics of the infiltrated pathogens, $p(t)$, is described (FIGURE S6) by

$$\frac{dp(t)}{dt} = \frac{k_4}{1+b_6 s(t)} - p(t)\left\{\frac{(1+d_4 p(t))(1+d_5 c_{17}(t))(1+d_6 c_{22}(t))(1+d_7 c_g(t))}{(1+b_7 c_4(t))(1+b_8 c_{13}(t))} + d_8\right\}, \qquad \text{(S5)}$$

where $k_4$ is the rate of pathogen infiltration, $d_4$, $d_5$, $d_6$, $d_7$, and $d_8$ are the elimination rates of infiltrated pathogens via infiltrated pathogens themselves, IL-17A, IL-22, IFNg (these factors increase release of AMPs as described below), and skin turnover, respectively, $b_6$ corresponds to the inhibitory strength for pathogens infiltration via skin barrier, $b_7$ and $b_8$ correspond to the inhibitory strengths for elimination of infiltrated pathogens via IL-4 and IL-13, respectively, and $c_g(t)$ is the concentration of IFNg. Levels of the infiltrated pathogens were defined between 0 (pathogen-free) and 1 (maximal infiltration).

We assumed that the steady-state infiltrated pathogen level is 1 (the maximal level of infiltrated pathogens when $\frac{dp(t)}{dt} = 0$) if skin barrier integrity is destructed completely ($s(t) = 0$) and the effects of AMP to decrease infiltrated pathogens is zero ($\frac{(1+d_4 p(t))(1+d_5 c_{17}(t))(1+d_6 c_{22}(t))(1+d_7 c_g(t))}{(1+b_7 c_4(t))(1+b_8 c_{13}(t))} \cong 0$) at the steady-state. This condition leads to $k_4 = d_8$.

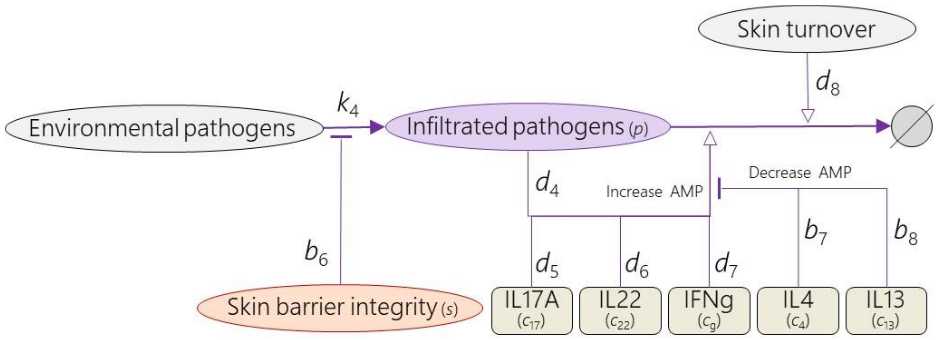


**FIGURE S6** Regulation of infiltrated pathogen level

The infiltrated pathogens increase by penetration of environmental pathogens through the skin barrier (with the rate $k_4$), whose integrity determines how easily pathogens can infiltrate (with strength $b_6$). Microbial pathogens are killed by AMP directly[46]. IL-17A[47], IL-22[48], and IFNg[49] increase AMP release from keratinocytes (with the strengths, $d_5$, $d_6$ and $d_7$, respectively), whereas the infiltrated pathogens, such as *Staphylococcus aureus*, also increase AMP release from keratinocytes via other pathways independent to these cytokines[50] ($d_4$). IL-4 and IL-13 inhibit the AMP release[49] (with the strengths $b_7$ and $b_8$). The infiltrated pathogens decrease due to the skin turnover ($d_8$).

#### 3.2. Cytokines

The dynamics of the cytokines and OX40L, $c_4(t)$, $c_{13}(t)$, $c_{17}(t)$, $c_{22}(t)$, $c_{31}(t)$, $c_{\mathrm{g}}(t)$, $c_{\mathrm{TS}}(t)$, and $c_{\mathrm{OX}}(t)$ are described (FIGURE S7) by

$$\frac{dc_4(t)}{dt} = k_{11}c_{\mathrm{t2}}(t) + k_{12} - d_{10}c_4(t), \quad \text{(S6)}$$
$$\frac{dc_{13}(t)}{dt} = k_{13}c_{\mathrm{t2}}(t) + k_{14} - d_{11}c_{13}(t), \quad \text{(S7)}$$
$$\frac{dc_{17}(t)}{dt} = k_{15}c_{\mathrm{t17}}(t) + k_{16} - d_{12}c_{17}(t), \quad \text{(S8)}$$
$$\frac{dc_{22}(t)}{dt} = k_{17}c_{\mathrm{t22}}(t) + k_{18} - d_{13}c_{22}(t), \quad \text{(S9)}$$
$$\frac{dc_{31}(t)}{dt} = k_{19}c_{\mathrm{t2}}(t) + k_{20} - d_{14}c_{31}(t), \quad \text{(S10)}$$
$$\frac{dc_{\mathrm{g}}(t)}{dt} = k_{21}c_{\mathrm{t1}}(t) + k_{22} - d_{15}c_{\mathrm{g}}(t), \quad \text{(S11)}$$
$$\frac{dc_{\mathrm{TS}}(t)}{dt} = k_{23}p(t) + k_{24} - d_{16}c_{\mathrm{TS}}(t), \quad \text{(S12)}$$
$$\frac{dc_{\mathrm{OX}}(t)}{dt} = k_{25}c_{\mathrm{TS}}(t) + k_{26} - d_{17}c_{\mathrm{OX}}(t), \quad \text{(S13)}$$

where $c_{\mathrm{TS}}(t)$ is the concentrations of TSLP and $c_{\mathrm{OX}}(t)$ is the level of OX40L, $k_{11}$ is the IL-4 secretion rate via Th2 , $k_{13}$ is the IL-13 secretion rate via Th2, $k_{15}$ is the IL-17A secretion rate via Th17, $k_{17}$ is the IL-22 secretion rate via Th22, $k_{19}$ is the IL-31 secretion rate via Th2, $k_{21}$ is are the IFNg secretion rate via Th1, $k_{23}$ is the TSLP secretion rate via infiltrated pathogens, $k_{25}$ is the OX40L expression rate via TSLP, $d_{10}$, $d_{11}$, $d_{12}$, $d_{13}$, $d_{14}$, $d_{15}$, $d_{16}$, and $d_{17}$ are the elimination rates for IL-4, IL-13, IL-17A, IL-22, IL-31, IFNg, TSLP, and OX40L, respectively, and $k_{12}$, $k_{14}$, $k_{16}$, $k_{18}$, $k_{20}$, $k_{22}$, $k_{24}$, and $k_{26}$ are the secretion or expression rates of IL-4, IL-13, IL-17A, IL-22, IL-31, IFNg, TSLP, and OX40L via other pathways that were not explicitly considered in the model.

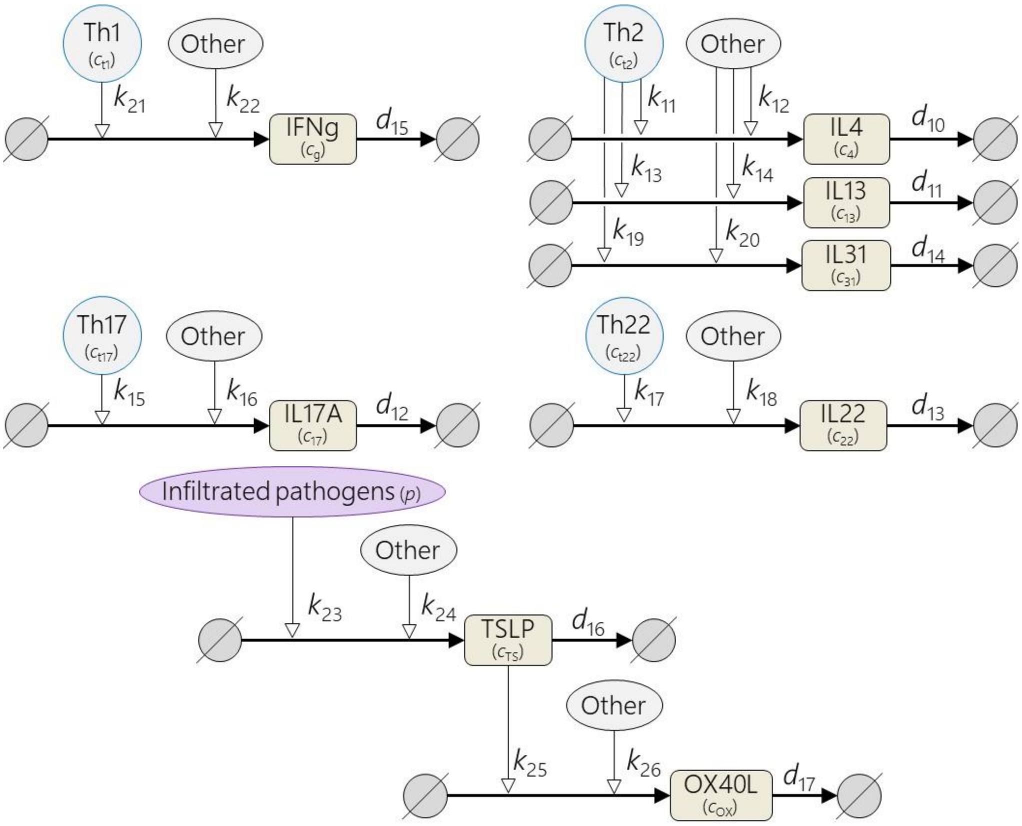


**FIGURE S7** Modelling of regulation of cytokine concentrations

Th1 secretes IFNg[51] (with the rate $k_{21}$), while Th2 secretes IL-4[52], IL-13[51], and IL31[53] (with the rates $k_{11}$, $k_{13}$, and $k_{19}$). Th17 and Th22 produce IL17A[52] and IL-22[54] (with the rates $k_{15}$ and $k_{17}$). Infiltrated pathogens induce TSLP in keratinocytes through TLR2 pathway[55] (with the rate $k_{23}$), and TSLP induces DCs to express OX40L[56] (with the rate $k_{26}$). There are other pathways releasing cytokines, which were implicitly described as “other” effects (with the rates $k_{12}$, $k_{14}$, $k_{16}$, $k_{18}$, $k_{20}$, $k_{22}$, $k_{24}$, $k_{26}$).

#### 3.3. T cells

The dynamics of the concentrations of Th1, Th2, Th17, and Th22 cells, $c_{\mathrm{t1}}(t)$, $c_{\mathrm{t2}}(t)$, $c_{\mathrm{t17}}(t)$, and $c_{\mathrm{t22}}(t)$, are described (FIGURE S8) by

$$\frac{dc_{\mathrm{t1}(t)}}{dt} = k_5 p(t)\left(\frac{1+k_9 c_{\mathrm{g}}(t)}{4+k_9 c_{\mathrm{g}}(t)+k_{10} c_4(t)}\right) - \frac{d_9 c_{\mathrm{t1}}(t)}{1+b_9 c_{\mathrm{OX}}(t)}, \quad \text{(S14)}$$

$$\frac{dc_{\mathrm{t2}}(t)}{dt} = k_6 p(t)\left(\frac{1+k_{10} c_4(t)}{4+k_9 c_{\mathrm{g}}(t)+k_{10} c_4(t)}\right) - \frac{d_9 c_{\mathrm{t2}}(t)}{1+b_9 c_{\mathrm{OX}}(t)}, \quad \text{(S15)}$$

$$\frac{dc_{\mathrm{t17}}(t)}{dt} = k_7 p(t)\left(\frac{1}{4+k_9 c_{\mathrm{g}}(t)+k_{10} c_4(t)}\right) - \frac{d_9 c_{\mathrm{t17}}(t)}{1+b_9 c_{\mathrm{OX}}(t)}, \quad \text{(S16)}$$

$$\frac{dc_{\mathrm{t22}}(t)}{dt} = k_8 p(t)\left(\frac{1}{4+k_9 c_{\mathrm{g}}(t)+k_{10} c_4(t)}\right) - \frac{d_9 c_{\mathrm{t22}}(t)}{1+b_9 c_{\mathrm{OX}}(t)}, \quad \text{(S17)}$$

where $k_5$, $k_6$, $k_7$, and $k_8$ are the rates of differentiation of naïve T cells to Th1, Th2, Th17, and Th22, respectively, $k_9$ and $k_{10}$ are the strengths of polarization for Th1 and Th2 differentiation, respectively, $d_9$ is the T cell elimination rate, $b_9$ is the inhibitory strength for T cells elimination by OX40L.

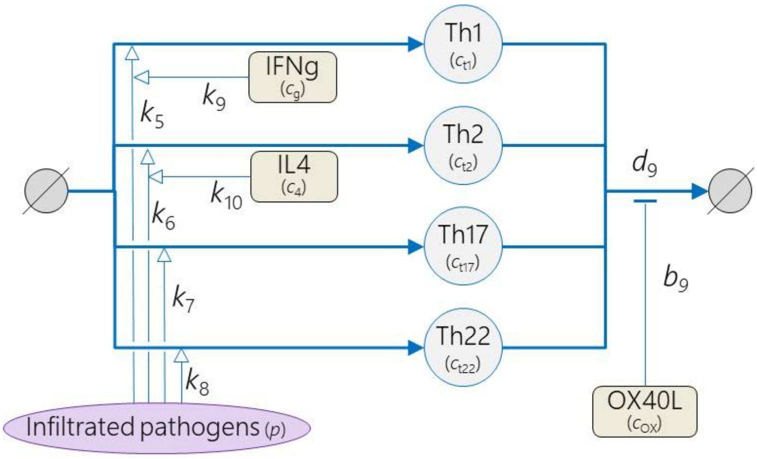


**FIGURE S8** Regulation of T-cell concentrations

Concentration of Th1, Th2, Th17, and Th22 cells increases via differentiation of naïve T cells[57] that are primed by dendritic cells (DCs) activated by the infiltrated pathogen[58] (with the rate $k_5$, $k_6$, $k_7$ and $k_8$, respectively). The balance of the differentiation into each T cell subset depends on the cytokine environment[59]; IFNg drives Th1 polarization[60] (with the strength $k_9$) while IL-4 promotes differentiation toward Th2[61] ($k_{10}$). Preferential polarization of certain helper-T cell subset reduces room for other subsets to be differentiated ($\mathrm{Th1} : \mathrm{Th2} : \mathrm{Th17} : \mathrm{Th22} = \left(1 + k_9 c_g(t)\right) : \left(1 + k_{10} c_4(t)\right) : 1 : 1$ with ratio to the sum of all the subtypes: $1 + k_9 c_g(t) + 1 + k_{10} c_4(t) + 1 + 1 = 4 + k_9 c_g(t) + k_{10} c_4(t)$). The T cells decrease according to their turnover (with the rate $d_9$). OX40L promotes prolongation of T cell activation and survival (with the strength $b_9$) as it activates the tumor necrosis factor receptor OX40, and functions as a T-cell costimulatory factor[62].

#### 3.4. Drug effects

We described the effects of the antibodies that inhibit the target signaling by scaling the actual concentrations of the target cytokines or OX40L by an inhibitory rate, $r_{\mathrm{inhibit}}$, for each drug (Eq. 2 and TABLE S3).

$r_{\mathrm{inhibit}}$ of each drug was defined using the half maximal inhibitory concentration ($\mathrm{IC}_{50}$) of the drug against the target biological factor and the mean concentration of drugs in the skin[63] ($d_{\mathrm{skin}}$) by

$$r_{\mathrm{inhibit}} = \begin{cases} \frac{d_{\mathrm{skin}}}{\mathrm{IC}_{50} + d_{\mathrm{skin}}} e_{\mathrm{a2}}, & \text{if drug is tralokinumab}, \\ \frac{d_{\mathrm{skin}}}{\mathrm{IC}_{50} + d_{\mathrm{skin}}}, & \text{otherwise}, \end{cases} \qquad \text{(S18)}$$

where $e_{\mathrm{a2}}$ represents the inhibitory effects of IL-13 binding to IL-13Rα2 in tralokinumab. $d_{\mathrm{skin}}$ is defined by $r_{\mathrm{skin/serum}} d_{\mathrm{serum}}$, where $r_{\mathrm{skin/serum}}$ is a ratio of drug concentration in the skin to that in serum and $d_{\mathrm{serum}}$ is the mean concentration of the drug in serum. We adopted $r_{\mathrm{skin/serum}} = 0.157$ for all the antibodies based on the estimated ratio of antibody concentration in the skin to that in the plasma[64]. Values of $\mathrm{IC}_{50}$ and $d_{\mathrm{serum}}$ (TABLE S3) were obtained from reported results of in vitro assay and the reported pharmacokinetics data of the adopted dose regimen (TABLE 1) in clinical trials.

We introduced $e_{\mathrm{a2}}$ for tralokinumab because tralokinumab not only inhibits IL-13 signaling via IL-13Rα1 but also enhances IL-13 signaling via inhibition of IL-13 binding to IL-13Rα2. IL-13Rα1 forms a heterodimeric receptor with IL-4Rα and is related to the effects of IL-13 signaling in AD pathogenesis. IL-13Rα2 is a decoy receptor to decrease IL-13 signaling via IL-13Rα1. Hence, the influence of the inhibition of IL-13Rα2 in tralokinumab was described by $e_{\mathrm{a2}}$ to scale down the $r_{\mathrm{inhibit}}$ based on the $\mathrm{IC}_{50}$ against IL-13Rα1. On the other hand, lebrikizumab prevents IL-13 from binding to IL-13Rα1 only[65]. We adopted $e_{\mathrm{a2}} = 0.44$ so that it reproduces the differences of %improved EASI and EASI-75 between tralokinumab and lebrikizumab (FIGURE 3).

We also described the effects of rIFNg that replaces the target cytokine (IFNg) by estimating the mean concentration, $c_{\mathrm{rIFNg}}$, of rIFNg in the skin (TABLE S3). $c_{\mathrm{rIFNg}}$ was estimated using the reported mean concentration of IFNg in the serum after rIFNg administration and the estimated ratio of rIFNg concentration in the skin to that in the plasma[66] by

$$c_{\mathrm{rIFNg}} = \frac{r_{\mathrm{IFNg\_skin/serum}} d_{\mathrm{rIFNg\_serum}}}{d_{\mathrm{IFNg\_skin}}}, \tag{S19}$$

where $d_{\mathrm{rIFNg\_serum}}$ is the mean concentration of rIFNg in serum after the rIFNg administration (i.e., the increased concentration of IFNg), $r_{\mathrm{IFNg\_skin/serum}}$ and $d_{\mathrm{IFNg\_skin}}$ are a ratio of rIFNg concentration in the skin to that in serum and mean concentration of IFNg in the skin in the observational study, respectively. $d_{\mathrm{rIFNg\_serum}}$ and $d_{\mathrm{IFNg\_skin}}$ were obtained from the pharmacokinetics data of rIFNg[67] and the IFNg concentration in the healthy skin in the observational study[68], respectively. We adopted $r_{\mathrm{IFNg\_skin/serum}} = 1.60$ based on the estimated ratio of macromolecule concentration in the skin to that in the plasma[66].

**TABLE S3** Parameter values for effects of drugs used in this study

| Drugs | $\mathrm{IC}_{50}$ (mcg/mL) | $d_{\mathrm{skin}}/d_{\mathrm{serum}}$ or $d_{\mathrm{rIFNg\_skin}}/d_{\mathrm{rIFNg\_serum}}$ (mcg/mL) | $r_{\mathrm{inhibit}}$ | | | | | | | $c_{\mathrm{rIFNg}}$ (fold change against healthy skin) |
|---|---|---|---|---|---|---|---|---|---|---|
| | | | IL-4 | IL-13 | IL-17A | IL-22 | IL-31 | TSLP | OX40L | IFNg |
| Dupilumab[69, 70] | IL-4: <0.01<br>IL-13: 0.01 | 183/29 | 0.99 | 0.99 | - | - | - | - | - | - |
| Nemolizumab[71,72] | 0.01 | 6/1 | - | - | - | - | 0.99 | - | - | - |
| Tezepelumab[73, 74] | 0.21 | 109/17 | - | - | - | - | - | 0.99 | - | - |
| GBR 830[75, 76] | 0.13 | 90/14 | - | - | - | - | - | - | 0.99 | - |
| Fezakinumab[a] | - | - | - | - | - | 0.99 | - | - | - | - |
| Secukinumab[77,78] | 0.08 | 45/7 | - | - | 0.99 | - | - | - | - | - |
| Tralokinumab[65, 79] | 0.10 | 70/11 | - | $0.99e_{\mathrm{a2}}$ | - | - | - | - | - | - |
| Lebrikizumab[80 b] | 0.10 | 91/14 | - | 0.99 | - | - | - | - | - | - |
| rIFNg[67] | - | 4.0E-6<br>/5.3E-4 | - | - | - | - | - | - | - | 2.1E2 |

a: $\mathrm{IC}_{50}$ and mean concentration were not found in published literature. We assumed an almost complete inhibition (the 99% inhibition as the same as other antibodies) for fezakinumab. b: $\mathrm{IC}_{50}$ and mean concentration were not found in the publication. We assumed that tralokinumab and lebrikizumab have the equivalent $\mathrm{IC}_{50}$ because their mechanisms of binding are similar[65] and that the mean drug concentration of 250 mg q2w is four times larger than that of 125 mg q4w (reported mean drug concentration is 22.8 mcg/mL).

### 4. Model parameters

We selected 102 parameters of the distributions ($\mu_n$ and $\sigma_n$ that define the distributions of the 51 parameters, TABLE S4) by

1) calculating $\mu_n$ of elimination rates for 11 biological factors based on available data and assumptions, and

2) tuning the remaining 91 parameter values so that the model reproduces clinical data.

**TABLE S4** Optimal parameter values obtained by parameter tuning

| Parameters | | $\mu_n$ | $\sigma_n$ | Comments |
|---|---|---|---|---|
| $k_1$ | Recovery rate of skin barrier integrity via skin turnover | 0.54 | 0.79 | - |
| $k_2$ | Recovery rate of skin barrier integrity via IL-22 | -1.50 | 0.39 | - |
| $k_3$ | Recovery rate of skin barrier integrity via placebo effects | 2.95 | 1.42 | - |
| $k_4$ | Rate of pathogen infiltration | - | - | $k_4 = d_8$ (Section S3.1 (b)) |
| $k_5$ | Rate of differentiation of naïve T cells to Th1 | 2.95 | 0.04 | - |
| $k_6$ | Rate of differentiation of naïve T cells to Th2 | 3.89 | 0.00 | - |
| $k_7$ | Rate of differentiation of naïve T cells to Th17 | 2.60 | 0.07 | - |
| $k_8$ | Rate of differentiation of naïve T cells to Th22 | 4.83 | 0.48 | - |
| $k_9$ | Strength of polarization for Th1 differentiation | -3.32 | 0.99 | - |
| $k_{10}$ | Strength of polarization for Th2 differentiation | -5.82 | 0.07 | - |
| $k_{11}$ | IL-4 secretion rate via Th2 | 5.50 | 0.16 | - |
| $k_{12}$ | IL-4 secretion rate via other pathways | 9.25 | 0.51 | - |
| $k_{13}$ | IL-13 secretion rate via Th2 | 6.65 | 0.37 | - |
| $k_{14}$ | IL-13 secretion rate via other pathways | 8.84 | 0.45 | - |
| $k_{15}$ | IL-17A secretion rate via Th17 | 4.01 | 0.27 | - |
| $k_{16}$ | IL-17A secretion rate via other factors | 2.64 | 0.85 | - |
| $k_{17}$ | IL-22 secretion rate via Th22 | 1.19 | 0.21 | - |
| $k_{18}$ | IL-22 secretion rate via other factors | 1.46 | 0.03 | - |
| $k_{19}$ | IL-31 secretion rate via Th2 | 1.54 | 0.19 | - |
| $k_{20}$ | IL-31 secretion rate via other factors | 1.99 | 0.58 | - |
| $k_{21}$ | IFNg secretion rate via Th1 | 0.46 | 1.00 | - |
| $k_{22}$ | IFNg secretion rate via other factors | 2.62 | 0.35 | - |
| $k_{23}$ | TSLP secretion rate via infiltrated pathogens | 4.00 | 0.52 | - |
| $k_{24}$ | TSLP secretion rate via other factors | 4.43 | 0.65 | - |
| $k_{25}$ | OX40L expression rates via TSLP | 0.42 | 0.48 | - |
| $k_{26}$ | OX40L expression rates via other factors | 1.68 | 0.78 | - |
| $b_1$ | Inhibitory strength for recovery of skin barrier via IL-4 | -8.69 | 0.57 | - |
| $b_2$ | Inhibitory strength for recovery of skin barrier via IL-13 | -3.54 | 1.58 | - |
| $b_3$ | Inhibitory strength for recovery of skin barrier via IL-17 | -3.92 | 0.35 | - |
| $b_4$ | Inhibitory strength for recovery of skin barrier via IL-22 | -0.59 | 0.80 | - |
| $b_5$ | Inhibitory strength for recovery of skin barrier via IL-31 | -2.09 | 0.90 | - |
| $b_6$ | inhibitory strength for pathogens infiltration via skin barrier | 0.39 | 0.44 | - |
| $b_7$ | Inhibitory strength for elimination of infiltrated pathogens via IL-4 | -7.99 | 0.09 | - |
| $b_8$ | Inhibitory strength for elimination of infiltrated pathogens via IL-13 | -3.50 | 0.05 | - |
| $b_9$ | Inhibitory strength for T cells elimination by OX40L | -2.66 | 0.30 | - |
| $d_1$ | Degradation rate of skin barrier via skin turnover | -1.60 | 1.53 | $\mu_n$ based on half-live (TABLE S5) |
| $d_2$ | Degradation rate of skin barrier via IL-31 | -2.64 | 0.57 | - |
| $d_3$ | Degradation rate of skin barrier via infiltrated pathogens | 1.25 | 2.28 | - |
| $d_4$ | Elimination rate of infiltrated pathogens via infiltrated pathogens themselves | -1.00 | 0.47 | - |
| $d_5$ | Elimination rate of infiltrated pathogens via IL-17A | -5.10 | 0.45 | - |
| $d_6$ | Elimination rate of infiltrated pathogens via IL-22 | -5.01 | 0.39 | - |
| $d_7$ | Elimination rate of infiltrated pathogens via IFNg | -8.70 | 0.62 | - |
| $d_8$ | Elimination rate of infiltrated pathogens via skin turnover | -1.60 | 0.04 | $\mu_n$ based on half-live (TABLE S5) |
| $d_9$ | T cell elimination rate | -0.55 | 0.09 | $\mu_n$ based on half-live (TABLE S5) |
| $d_{10}$ | Elimination rates for IL-4 | 5.91 | 0.22 | $\mu_n$ based on half-live (TABLE S5) |
| $d_{11}$ | Elimination rates for IL-13 | 5.91 | 0.11 | $\mu_n$ based on half-live (TABLE S5) |
| $d_{12}$ | Elimination rates for IL-17A | 3.33 | 0.57 | $\mu_n$ based on half-live (TABLE S5) |
| $d_{13}$ | Elimination rates for IL-22 | 3.33 | 0.56 | $\mu_n$ based on half-live (TABLE S5) |
| $d_{14}$ | Elimination rates for IL-31 | 3.33 | 0.10 | $\mu_n$ based on half-live (TABLE S5) |
| $d_{15}$ | Elimination rates for IFNg | 2.68 | 0.15 | $\mu_n$ based on half-live (TABLE S5) |
| $d_{16}$ | Elimination rates for TSLP | 3.33 | 0.20 | $\mu_n$ based on half-live (TABLE S5) |
| $d_{17}$ | Elimination rates for OX40L | 0.48 | 0.24 | $\mu_n$ based on half-live (TABLE S5) |

### 4.1. $\mu_n$ of elimination rates calculated with published data and assumptions

The elimination rates of the 11 biological factors were determined using the half-lives measured in vivo (serum) in humans (TABLE S5). The calculated values were set as the $\mu_n$, the mean value for each distribution.

**TABLE S5** Elimination rates of biological factors used in this study

| Parameters | Biological factors | Elimination rate (/week) | Half-life (week) | Comments on half-lives |
|---|---|---|---|---|
| $d_1$ | Skin barrier integrity | 0.202 | 3.4[81] | Based on the half-life of the epidermis |
| $d_8$ | Infiltrated pathogens | 0.202 | 3.4[81] | Based on the half-life of the epidermis |
| $d_9$ | Th1 | 0.578 | 1.2[82] | - |
| | Th2 | | | |
| | Th17 | | | |
| | Th22 | | | |
| $d_{10}$ | IL-4 | 368 | 1.9E-03[83] | - |
| $d_{11}$ | IL-13 | 368 | 1.9E-03 | Assumed same as IL-4 because IL-4 and IL-13 are the same IL-2 sub-family |
| $d_{12}$ | IL-17A | 28.0 | 2.5E-02 | Assumed mean value of IL-4 and IFNg as average cytokine |
| $d_{13}$ | IL-22 | 28.0 | 2.5E-02 | Assumed mean value of IL-4 and IFNg as average cytokine |
| $d_{14}$ | IL-31 | 28.0 | 2.5E-02 | Assumed mean value of IL-4 and IFNg as average cytokine |
| $d_{15}$ | IFNg | 14.6 | 4.8E-02[84] | - |
| $d_{16}$ | TSLP | 28.0 | 2.5E-02 | Assumed mean value of IL-4 and IFNg as average cytokine |
| $d_{17}$ | OX40L | 1.62 | 0.4[85] | Based on dendritic cells of mouse data. OX40L are expressed on dendritic cells. |

### 4.2. Parameters tuned to reproduce clinical data

The remaining 91 parameters were tuned so that the model reproduces the following clinical data consisting of 136 reference values;

- mean values and coefficient of variation (%CV) of 12 biological factors (IL-4, IL-13, IL-17A, IL-22, IL-31, IFNg, TSLP, OX40L, Th1, Th2, Th17, and Th22) in observational studies and baseline EASI score in clinical trial of dupilumab (TABLE S2. 2 indices x 13 factors = 26 reference values) and
- mean EASI score and EASI-75 when 9 drugs and placebo were applied in clinical trials (FIGURE 1. 2 indices x 10 interventions x 1-10 time points/intervention = 110 reference values).

We searched the parameters that minimizes the cost function, ***J***, defined by

$$\boldsymbol{J} = w_1 J_1 + w_2 J_2 + w_3 J_3 + w_4 J_4, \tag{S18}$$

where

$$J_1 = \sqrt{\frac{1}{l}\sum_{h=1}^{l}(u_h - \hat{u}_h)^2}, \tag{S19}$$

$$J_2 = \sqrt{\frac{1}{l}\sum_{h=1}^{l}(v_h - \hat{v}_h)^2}, \tag{S20}$$

$$J_3 = \sqrt{\frac{1}{n}\sum_{i=1}^{n}\left\{\frac{1}{m}\sum_{j=1}^{m} w_{\mathrm{T}}\left(a_i(t_j) - \hat{a}_i(t_j)\right)\right\}^2}, \tag{S21}$$

$$J_4 = \sqrt{\frac{1}{n}\sum_{i=1}^{n}\left\{\frac{1}{m}\sum_{j=1}^{m} w_{\mathrm{T}}\left(b_i(t_j) - \hat{b}_i(t_j)\right)\right\}^2}. \tag{S22}$$

The terms, $J_1$ and $J_2$ are root mean squared errors of mean values and %CV of biological factors, respectively. $J_3$ and $J_4$ are weighted root mean squared errors of mean EASI score and EASI-75, respectively. $w_1$ to $w_4$ are the weighting coefficients. $u_h$ and $v_h$ are the mean value and the %CV of the *h*-th biological factor (*h*=1, …, *l*) in the observational studies. $\hat{u}_h$ and $\hat{v}_h$ are the corresponding values among 1000 virtual patients in the simulation at the steady-state (after 1000 weeks). $a_i(t_j)$ and $b_i(t_j)$ are the reference value of the mean EASI score and of EASI-75 in the study using the *i*-th drug (*i*=1, …, *n*) at time $t_j$ (*j*=1,…,*m*) with the weighting for time points, $w_{\mathrm{T}}$ . $\hat{a}_i(t_j)$ and $\hat{b}_i(t_j)$ are the corresponding simulated values. We used $(w_1, w_2, w_3, w_4) = (10,1,1,1)$ and $w_{\mathrm{T}} = \begin{cases} 1\ (t_j < 8) \\ 10\ (t_j \geq 8) \end{cases}$. We set a higher weight on the mean values of the biological factors, $w_1$, because its fitting error tends to be smaller than other items (ie, %CV of the biological factors, the mean EASI score, and EASI-75). We also set a higher weight on the time points of 8 weeks or later, $w_{\mathrm{T}}$, because the efficacies are evaluated at least after 8 weeks as primary endpoints in the clinical trials[86].

Parameter tuning was based on differential evolution[87], which is known to be an effective method for global optimization of a large number of parameters. The conditions for differential evolution were set as follows based on manual trial-and-error.

| | |
|---|---|
| Mutation constant (F) | : 0.5 |
| Crossover constant (CR) | : 0.7 |
| Strategy | : DE/best/1/bin |
| Number of population vectors (NP) | : 92 |
| Number of function evaluations (nfe) | : 46092 |
| Ranges of parameters searched | : TABLE S6 |

We tuned both the 91 parameters of the distributions and $e_{a2}$ (92 parameters in total) at once. ***J*** was minimized to 27.1, where the model fitness was confirmed visually (FIGURE 3).
As reference information, we also calculated (not-weighted) root mean squared errors of mean EASI score and EASI-75 by

$$J_5 = \sqrt{\frac{1}{n}\sum_{i=1}^{n}\left\{\frac{1}{m}\sum_{j=1}^{m}\left(a_i(t_j) - \hat{a}_i(t_j)\right)\right\}^2}, \quad \text{(S23)}$$

$$J_6 = \sqrt{\frac{1}{n}\sum_{i=1}^{n}\left\{\frac{1}{m}\sum_{j=1}^{m}\left(b_i(t_j) - \hat{b}_i(t_j)\right)\right\}^2}. \quad \text{(S24)}$$

**TABLE S6** Ranges of parameters searched in parameter tuning

| Parameters | | $\mu_n$ | $\sigma_n$ | Comments |
|---|---|---|---|---|
| $k_1$ | Recovery rate of skin barrier integrity via skin turnover | [0, 1] | [0, 1] | - |
| $k_2$ | Recovery rate of skin barrier integrity via IL-22 | [-2, -1] | [0, 1] | - |
| $k_3$ | Recovery rate of skin barrier integrity via placebo effects | [2, 3] | [1, 2] | - |
| $k_4$ | Rate of pathogen infiltration | - | - | $k_4 = d_8$ (Section S3.1 (b)) |
| $k_5$ | Rate of differentiation of naïve T cells to Th1 | [2, 3] | [0, 1] | - |
| $k_6$ | Rate of differentiation of naïve T cells to Th2 | [3, 4] | [0, 1] | - |
| $k_7$ | Rate of differentiation of naïve T cells to Th17 | [2, 3] | [0, 1] | - |
| $k_8$ | Rate of differentiation of naïve T cells to Th22 | [4, 5] | [0, 1] | - |
| $k_9$ | Strength of polarization for Th1 differentiation | [-4, -3] | [0, 1] | - |
| $k_{10}$ | Strength of polarization for Th2 differentiation | [-6, -5] | [0, 1] | - |
| $k_{11}$ | IL-4 secretion rate via Th2 | [5, 6] | [0, 1] | - |
| $k_{12}$ | IL-4 secretion rate via other pathways | [9, 10] | [0, 1] | - |
| $k_{13}$ | IL-13 secretion rate via Th2 | [6, 7] | [0, 1] | - |
| $k_{14}$ | IL-13 secretion rate via other pathways | [8, 9] | [0, 1] | - |
| $k_{15}$ | IL-17A secretion rate via Th17 | [4, 5] | [0, 1] | - |
| $k_{16}$ | IL-17A secretion rate via other factors | [2, 3] | [0, 1] | - |
| $k_{17}$ | IL-22 secretion rate via Th22 | [1, 2] | [0, 1] | - |
| $k_{18}$ | IL-22 secretion rate via other factors | [1, 2] | [0, 1] | - |
| $k_{19}$ | IL-31 secretion rate via Th2 | [1, 2] | [0, 1] | - |
| $k_{20}$ | IL-31 secretion rate via other factors | [1, 2] | [0, 1] | - |
| $k_{21}$ | IFNg secretion rate via Th1 | [0, 1] | [1, 2] | - |
| $k_{22}$ | IFNg secretion rate via other factors | [2, 3] | [0, 1] | - |
| $k_{23}$ | TSLP secretion rate via infiltrated pathogens | [4, 5] | [0, 1] | - |
| $k_{24}$ | TSLP secretion rate via other factors | [4, 5] | [0, 1] | - |
| $k_{25}$ | OX40L expression rates via TSLP | [0, 1] | [0, 1] | - |
| $k_{26}$ | OX40L expression rates via other factors | [1, 2] | [0, 1] | - |
| $b_1$ | Inhibitory strength for recovery of skin barrier via IL-4 | [-9, -8] | [0, 1] | - |
| $b_2$ | Inhibitory strength for recovery of skin barrier via IL-13 | [-4, -3] | [1, 2] | - |
| $b_3$ | Inhibitory strength for recovery of skin barrier via IL-17 | [-4, -3] | [0, 1] | - |
| $b_4$ | Inhibitory strength for recovery of skin barrier via IL-22 | [-1, 0] | [0, 1] | - |
| $b_5$ | Inhibitory strength for recovery of skin barrier via IL-31 | [-3, -2] | [0, 1] | - |
| $b_6$ | inhibitory strength for pathogens infiltration via skin barrier | [0, 1] | [0, 1] | - |
| $b_7$ | Inhibitory strength for elimination of infiltrated pathogens via IL-4 | [-8, -7] | [0, 1] | - |
| $b_8$ | Inhibitory strength for elimination of infiltrated pathogens via IL-13 | [-4, -3] | [0, 1] | - |
| $b_9$ | Inhibitory strength for T cells elimination by OX40L | [-3, -2] | [0, 1] | - |
| $d_1$ | Degradation rate of skin barrier via skin turnover | - | [1, 2] | $\mu_n$ based on half-live (TABLE S5) |
| $d_2$ | Degradation rate of skin barrier via IL-31 | [-3, -2] | [0, 1] | |
| $d_3$ | Degradation rate of skin barrier via infiltrated pathogens | [1, 2] | [2, 3] | - |
| $d_4$ | Elimination rate of infiltrated pathogens via infiltrated pathogens themselves | [-2, -1] | [0, 1] | - |
| $d_5$ | Elimination rate of infiltrated pathogens via IL-17A | [-6, -5] | [0, 1] | - |
| $d_6$ | Elimination rate of infiltrated pathogens via IL-22 | [-6, -5] | [0, 1] | - |
| $d_7$ | Elimination rate of infiltrated pathogens via IFNg | [-9, -8] | [0, 1] | - |
| $d_8$ | Elimination rate of infiltrated pathogens via skin turnover | - | [0, 1] | $\mu_n$ based on half-live (TABLE S5) |
| $d_9$ | T cell elimination rate | - | [0, 1] | $\mu_n$ based on half-live (TABLE S5) |
| $d_{10}$ | Elimination rates for IL-4 | - | [0, 1] | $\mu_n$ based on half-live (TABLE S5) |
| $d_{11}$ | Elimination rates for IL-13 | - | [0, 1] | $\mu_n$ based on half-live (TABLE S5) |
| $d_{12}$ | Elimination rates for IL-17A | - | [0, 1] | $\mu_n$ based on half-live (TABLE S5) |
| $d_{13}$ | Elimination rates for IL-22 | - | [0, 1] | $\mu_n$ based on half-live (TABLE S5) |
| $d_{14}$ | Elimination rates for IL-31 | - | [0, 1] | $\mu_n$ based on half-live (TABLE S5) |
| $d_{15}$ | Elimination rates for IFNg | - | [0, 1] | $\mu_n$ based on half-live (TABLE S5) |
| $d_{16}$ | Elimination rates for TSLP | - | [0, 1] | $\mu_n$ based on half-live (TABLE S5) |
| $d_{17}$ | Elimination rates for OX40L | - | [0, 1] | $\mu_n$ based on half-live (TABLE S5) |

### 5. Influences of pathophysiological backgrounds of virtual patients on clinical efficacy

We investigated the influence of the 51 model parameters on %improve EASI of each drug using the LHS-PRCC (FIGURE 4), and simulated the %improve EASI at week 24 after patient stratification based on the baseline levels of cytokines in the skin (FIGURE S9) with regards to drugs other than dupilumab as well.

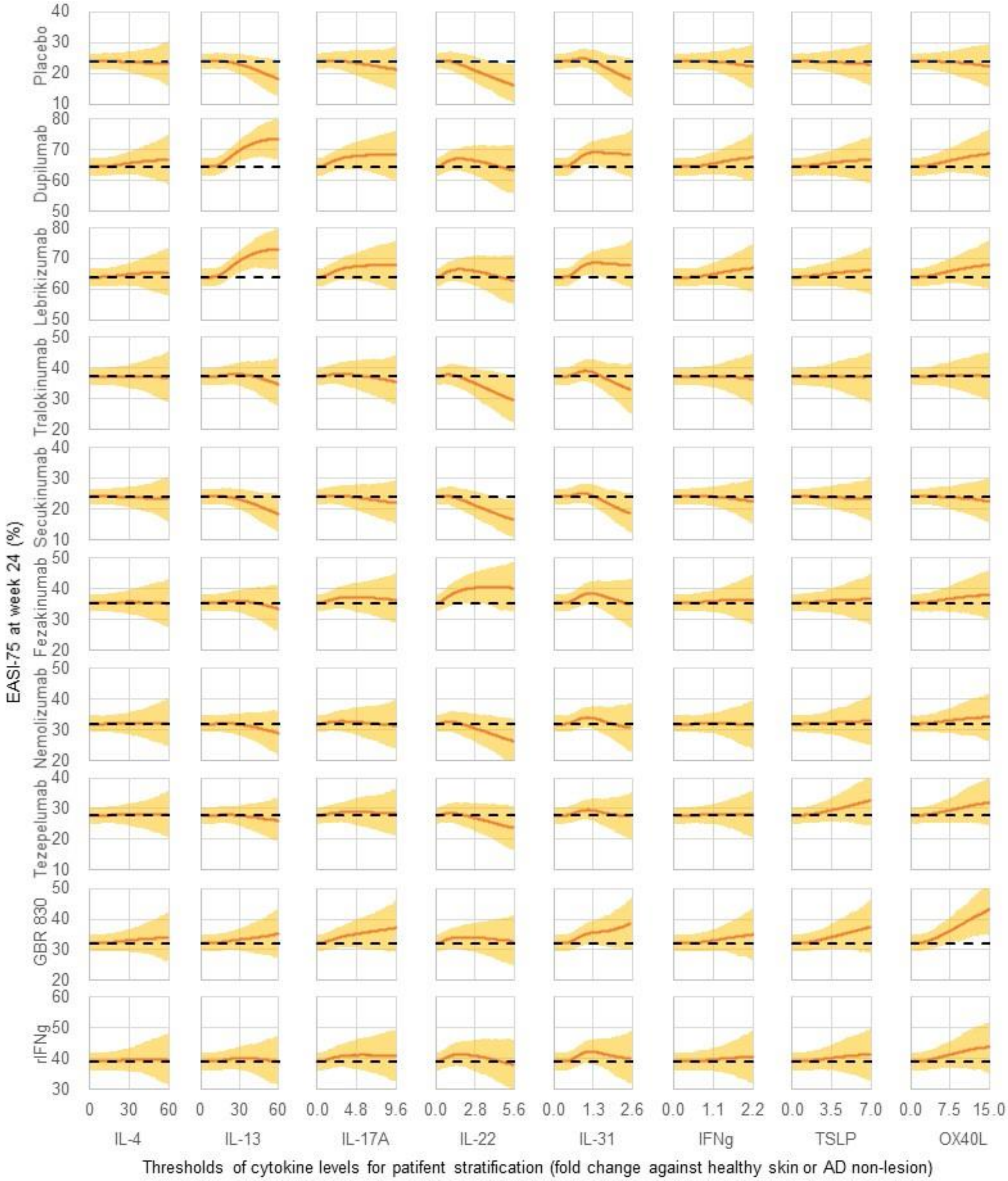


**FIGURE S9** Simulated EASI-75 at 24 weeks after drug treatment (y-axis) for stratified patients with their cytokine baseline levels larger than the threshold values (x-axis)

The number of stratified virtual patients decreases for a larger threshold value. The threshold of zero includes all the virtual patients and the maximum threshold value corresponds to inclusion of at least 10% of 1000 virtual patients, who were generated according to the tuned distributions of the parameters (TABLE S4). Simulation was iterated 1000 times where a cohort of 1000 virtual patients were created at each iteration. Lines and shaded areas are the mean value and 95% CI of 1000 simulations, respectively. The higher EASI-75 compared with that without patient stratification (with the threshold value of zero: dashed line) suggests a success in stratifying good responders.

#### 5.1. Influence of skin barrier on placebo effects

Three skin barrier-related parameters ($k_1$, $k_3$, and $b_6$) had a significant PRCC with the efficacy in placebo group (FIGURE 4); these influences were also observed in all the drugs as the placebo effects were considered in both placebo- and drug-treated groups in the simulation.

The virtual patients with a lower $k_1$ and a higher $k_3$ benefited more from recovery of skin barrier via placebo effects, and thereby achieved a higher %improve EASI. The virtual patients with a higher $b_6$ inhibit pathogen infiltration more strongly through the recovery of skin barrier by placebo effects, and thereby achieved higher %improve EASI. These results may suggest that older patients whose skin has a slow baseline turnover (a lower $k_1$) but can inhibit pathogen infiltration (a higher $b_6$, e.g., sufficient filaggrin functions to form skin barrier) are more responsive to placebo effects.

#### 5.2. Influence of skin barrier on efficacy in multiple drugs in common

Four skin barrier-related parameters ($b_2$, $b_4$, $d_1$, and $d_3$) had a significant PRCC with the %improved EASI by several drugs (FIGURE 4). These influences correspond to baseline severity of skin barrier defects rather than MoA of each drug.

Virtual patients with higher $b_2$, $b_4$, $d_1$, and $d_3$ were more responsive to treatment by several drugs (i.e., dupilumab, lebrikizumab, fezakinumab, and rIFNg) because they benefit from recovery of skin barrier via each drug MoA, achieving a higher %improve EASI. These results suggest that patients with a higher degradation rate of skin barrier via skin turnover ($d_1$, e.g., activity of kallikreins to degrade skin barrier) and via infiltrated pathogens ($d_3$, e.g., amounts of extracellular protease from S. aureus to degrade skin barrier), as well as a larger influence of cytokines on skin barrier damage ($b_2$ and $b_4$ correspond to IL-13 and IL-22), are more responsive to several drugs.

#### 5.3. Influence of IL-13 level on efficacy in lebrikizumab and tralokinumab

The results for lebrikizumab and tralokinumab were similar to those about dupilumab as both drugs have the drug target IL-13 in common, although tralokinumab was less influenced by the IL-13-related parameters due to the lower inhibition rate of IL-13 than lebrikizumab (estimated 44% of lebrikizumab: $e_{\mathrm{a2}} = 0.44$).

Ten model parameters had a significant PRCC with the %improved EASI by lebrikizumab (FIGURE 4). Four out of the ten parameters are IL-13-related ($k_{13}$, $k_{14}$, $b_2$, and $d_{11}$), and the remaining six parameters are skin barrier-related parameters ($k_1$, $k_3$, $b_4$, $b_6$ $d_1$, and $d_3$) that correspond to placebo effects and baseline severity of skin barrier defects rather than MoA of lebrikizumab.

The four IL-13 related parameters ($k_{13}$, $k_{14}$, $b_2$, and $d_{11}$) characterize responders for lebrikizumab. Virtual patients with higher $k_{13}$, $k_{14}$, and $b_2$ and a lower $d_{11}$ were more responsive to treatment by lebrikizumab. The parameters, $k_{13}$, $k_{14}$, and $d_{11}$, affect the IL-13 baseline level, and EASI-75 was improved by stratifying virtual patients with a higher IL-13 baseline level (FIGURE S9). The parameter, $b_2$, describes the influence of IL-13 on skin barrier damage.

Four model parameters had a significant PRCC with the %improved EASI by tralokinumab (FIGURE 4). One out of the six parameters are IL-13-related ($b_2$), and the remaining five parameters are skin barrier-related parameters ($k_1$, $k_3$, and $b_6$) that correspond to placebo effects rather than MoA of tralokinumab.

Virtual patients with higher $b_2$ were more responsive to treatment by tralokinumab, although the extent was smaller than the case of lebrikizumab. As opposed to lebrikizumab, EASI-75 was not improved by stratifying virtual patients with a higher IL-13 baseline level (FIGURE S8).

#### 5.4. Influence of IL-22 level on efficacy in fezakinumab

Eight model parameters had a significant PRCC with the %improved EASI by fezakinumab (FIGURE 4). Two out of the eight parameters are IL-22-related ($b_4$ and $d_{13}$), and the remaining six parameters are skin barrier-related parameters ($k_1$, $k_3$, $b_2$, $b_6$ $d_1$, and $d_3$) that correspond to placebo effects and baseline severity of skin barrier defects rather than MoA of fezakinumab.

The two IL-22-related parameters ($b_4$ and $d_{13}$) can characterize responders for fezakinumab as virtual patients with lower $d_{13}$ were more responsive to treatment by fezakinumab. The parameter, $d_{13}$, affect the IL-22 baseline level, and EASI-75 was slightly improved by stratifying virtual patients with a higher IL-22 baseline level (FIGURE S9). It is consistent with the results from actual clinical trials of fezakinumab, where a higher efficacy was observed in the AD patients with higher baseline mRNA levels of IL-22[88]. The parameter, $b_4$, describes the influence of IL-22 on skin barrier damage.

#### 5.5. Influence of Th1 polarization on efficacy in rIFNg

Eight model parameters had a significant PRCC with the %improved EASI by rIFNg (FIGURE 4). One out of the nine parameters is strength of polarization for Th1 differentiation ($k_9$), which is related to IFNg (i.e., IFNg drives Th1 polarization), and the remaining seven parameters are skin barrier-related parameters ($k_1$, $k_3$, $b_2$, $b_4$, $b_6$, $d_1$, and $d_3$) that correspond to placebo effects and baseline severity of skin barrier defects rather than MoA of rIFNg. There was no improvement in EASI-75 by stratifying virtual patients with any cytokine baseline level (FIGURE S9).

### 6. **Comparison of the parameter distributions between dupilumab good and poor responders**

We compared distributions of the model parameters between good and poor responders for dupilumab based on the EASI-75 criterion (FIGURE S10), and confirmed that the ten parameters ($k_1$, $k_3$, $k_{13}$, $k_{14}$, $b_2$, $b_4$, $b_6$, $d_1$, $d_3$, and $d_{11}$) that demonstrated a significant influence on %improve EASI of dupilumab (FIGURE 4) had different distributions for good and poor responders for dupilumab. Among the ten parameters, three parameters with larger PRCC ($k_1$: -0.6, $k_3$: 0.6, and $b_2$: 0.6) with %improve EASI of dupilumab showed a larger difference in distributions between dupilumab good and poor responders, compared to the other seven parameters that has a smaller PRCC ($k_{13}$, $k_{14}$, $b_4$, $b_6$, $d_1$, $d_3$, and $d_{11}$: -0.2 ~ 0.3).

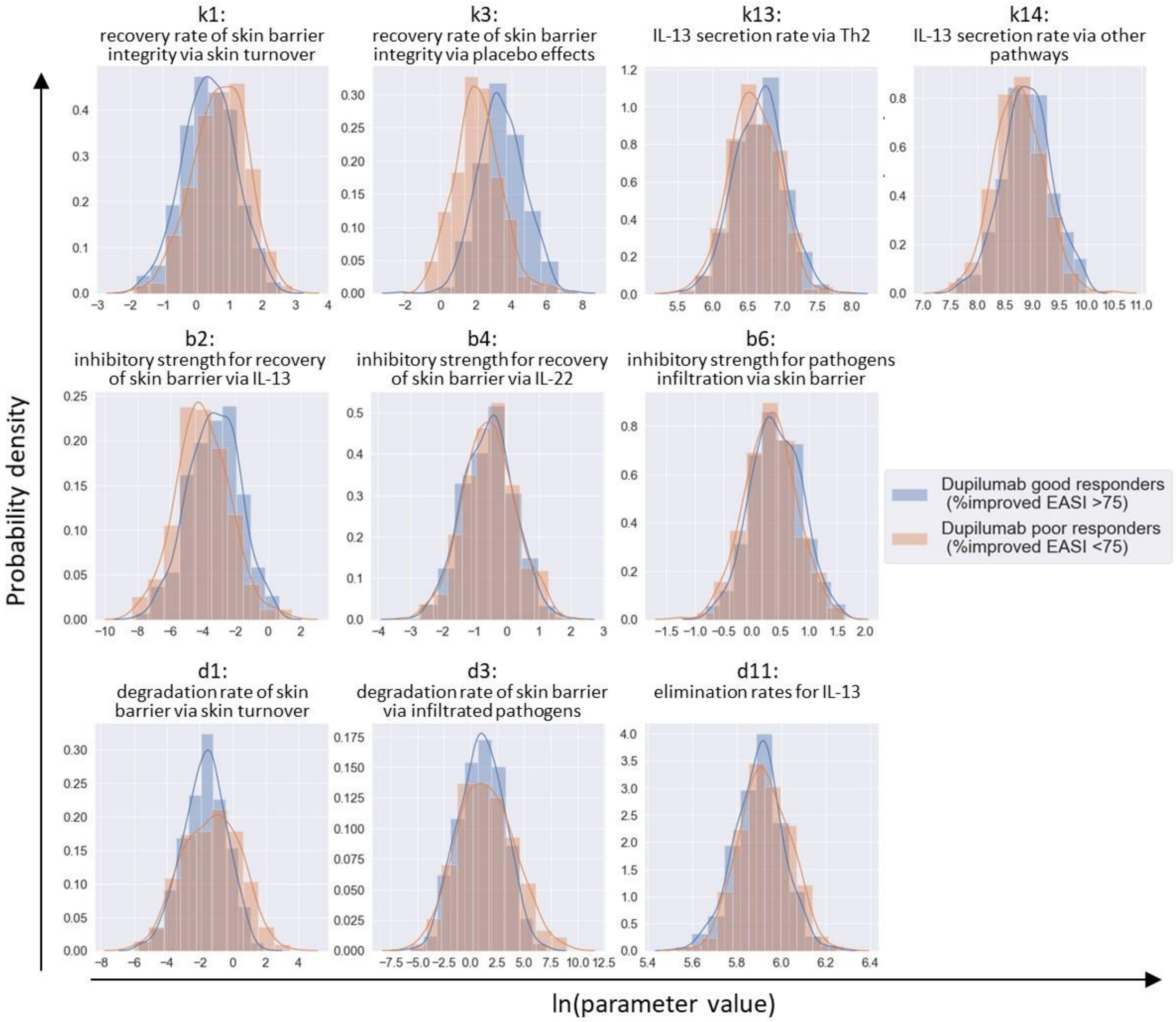


**FIGURE S10** Distributions of model parameters for dupilumab good (blue) and poor (orange) responders.

The comparison visually confirmed that the 11 parameters ($k_1$, $k_3$, $k_{13}$, $k_{14}$, $b_2$, $b_4$, $b_6$, $d_1$, $d_3$, and $d_{11}$) that had significant influence on %improve EASI of dupilumab (FIGURE 4) were differently distributed between dupilumab good and poor responders. We generated a large number of virtual patients (1000 virtual patients = 1000 sets of 52 parameter values) by randomly sampling each of the parameter values from the distribution in Eq. (3). The virtual patients were treated with dupilumab to simulate %improved EASI at 24 weeks and were categorized into dupilumab good responders (%improved EASI>75) or dupilumab poor responders (%improved EASI<75). Distributions of the 11 parameters were plotted for dupilumab good and poor responders separately.
